# Lifestyle and BrainAGE in Adult Depression

**DOI:** 10.1101/2025.03.27.25324698

**Authors:** Nicole Sanford, Yuetong Yu, Ruiyang Ge, Shalaila S. Haas, Liisa A.M. Galea, Sophia Frangou

## Abstract

Background: This study tested whether lifestyle and fitness features that influence brain health in the general population differentially affect adults with a history of depression. Brain health was assessed using the brain-age-gap-estimate (brainAGE), a personalized index of the brain’s biological age. Methods: Medically healthy adults (44-82 years) from the UK Biobank with a history of depression (n=896) or no psychiatric history (n=36,206) were included. Heterogeneity Through Discriminative Analysis was used to cluster the depression group based on 224 lifestyle and fitness features. Global and local (voxel-based) brainAGE were computed from structural neuroimaging data. The study design and implementation involved input from the position of related lived experience. Outcomes: Four depression clusters were identified. The “balanced moderates” cluster (n=253) had good health and balanced lifestyle habits. The “optimal diet and activity” cluster (n=178) had good health, healthy diets, and regular physical activity. The “metabolic risk-sedentary” cluster (n=315) had higher body mass index, poor diet, and sedentary behaviour. The “frailty-low activity” cluster (n=150) had a varied diet coupled with physical frailty. Mood symptoms were lowest in the “balanced moderates” cluster and highest in the “metabolic risk-sedentary” cluster. The presence of a history of depression was associated with older global brainAGE and with local brainAGE in ventromedial prefrontal regions regardless of cluster assignment. The “metabolic risk-sedentary” cluster also exhibited elevated local brainAGE in the hippocampal complex and thalamus. Interpretation: This study highlights the heterogeneity in lifestyle and fitness factors among adults with a history of depression, underscoring the detrimental influence of depression as well as poor diet and physical inactivity on the biological age of the brain.

## INTRODUCTION

Major depressive disorder (MDD) is intricately linked to brain health,^1,2^ making brain health a critical factor in managing its treatment. In recent years, the brain-age-gap estimate (brainAGE), which is an individualized measure of the pace of the biological aging of the brain, has emerged as a robust indicator of brain health.^3,4^ Patients with MDD have higher brainAGE compared to healthy individuals,^5,6^ which has been associated with poor antidepressant response in some but not all studies,^7,8^ poor working memory,^9,10^ increased symptomatic severity,^11^ and greater disability.^9^

Higher brainAGE in MDD has been associated with amplified inflammatory responses during depressive episodes which may lead to accelerated cellular aging.^12,13^ However, lifestyle factors that contribute to older brainAGE in the general population,^14,15^ notably social support, sleep, physical exercise, and healthy dietary intake, may also be important for brain health in the context of MDD. Support for this notion is based on extensive literature linking these same lifestyle factors to the risk of MDD.^16^ By contrast, a protective role has been proposed for physical activity^17,18^ and for healthy calorie daily intake based on fruits, vegetables, whole grains, lean proteins, and low-fat dairy products while limiting added sugars and saturated fats.^18,19^ These findings highlight the importance of investigating how brainAGE and psychopathology in individuals with a history of depression may be differentially affected by distinct lifestyle factors and physical traits that influence both the pace of aging and the risk of depression.

To address this issue, the current study leveraged the rich dataset of the UK Biobank (UKB) using neuroimaging data and detailed information on health and lifestyle.^20^ The analytical design benefits from two innovative elements. First, we used Heterogeneity Through Discriminative Analysis (HYDRA)^21^ to identify clusters of UKB participants with a history of depression based on lifestyle factors and physical traits relating to general health. Our rationale for this analysis was based on prior evidence that beneficial and unhealthy lifestyle choices tend to cluster within individuals and are not distributed uniformly in the population.^22–24^ Second, in addition to global brainAGE (G-brainAGE), which is computed based on global age-related brain changes, we also calculated local brainAGE (L-brainAGE) based on a pre-trained U-Net model^25^ to yield fine-grained spatial information on the anatomical patterns of brain aging in our study sample.

Our working hypotheses were that individuals with a history of depression (a) would exhibit an older brainAGE compared to psychiatrically healthy individuals, and (b) that this association would be more pronounced among those with lifestyle factors and physical traits known to confer greater risk for older brainAGE.

## METHODS

### Sample

The UKB is a population-based cohort study of over 500,000 self-referred individuals from the United Kingdom assessed at one of 22 centres in Scotland, England, and Wales between 2006 and 2010.^20^ In addition to T1-weighted structural magnetic resonance imaging (MRI), detailed information on health and lifestyle was collected for each participant. The UKB has ethical approval from the Northwest Multi-Centre Research Ethics Committee (MREC). All participants provided informed consent. Further details on the UKB Ethics and Governance framework are available here: https://www.ukbiobank.ac.uk/learn-more-about-uk-biobank/governance. The present analyses were conducted under data application number 69022.

A total of 37,102 participants were selected for the current study based on the following eligibility criteria. Participants were excluded from the present study if they rated their overall health as “poor” (UKB field 2178), had a history of stroke (field 4056), had a known history of any diseases of the nervous system as per ICD-10 diagnoses acquired at their first assessment (field 41270, ICD Chapter VI), or had neuroimaging data of poor quality and/or preprocessing errors (Supplemental Figure S1). Following the above exclusions, participants with a history of at least one major depressive episode (field 41270, ICD Chapter V, ICD codes F32-F33) comprised the depression group. We employed the ICD-10 classification for depressive episodes as it has been found to be an effective UKB depression criterion for identifying disease-relevant alterations in the brain.^26^ Participants without a history of psychiatric disorders as per their ICD-10 diagnoses (field 41270, ICD Chapter V, ICD codes F20-F39) formed the non-psychiatric comparison group. The final sample comprised 36,206 non-psychiatric comparison individuals (18,974 females; mean age = 63.84 years) and 896 individuals with a history of MDD (591 females; mean age = 63.10 years).

### Neuroimaging

The whole-brain T1-weighted structural MRI scans of the UKB sample were acquired on Siemens Skyra 3T scanners running VD13A SP4, with a standard Siemens 32-channel RF head coil using a 3D MPRAGE sequence with 1mm isotropic resolution and a 208 × 256 × 256 field-of-view (full acquisition details at https://biobank.ctsu.ox.ac.uk/crystal/crystal/docs/brain_mri.pdf). The participants’ NIFTI images were downloaded from the UKB repository and processed locally at the Advanced Research Computing Services at the University of British Columbia (UBC). For computing G-brainAGE, the images were processed using standard pipelines applied in FreeSurfer 7.1 (https://surfer.nmr.mgh.harvard.edu/fswiki/rel7downloads) to yield brain morphometric measures of intracranial volume, regional cortical thickness and surface area, and regional subcortical volumes. For computing L-brainAGE, the images were segmented into grey and white matter. The Diffeomorphic Anatomic Registration Through Exponentiated Lie algebra algorithm (DARTEL)^27^ was applied to normalize the segmented scans into a standard MNI space (MNI-152 space), followed by resampling to 1.5mm^3^ with a 4mm smoothing kernel.

G-brainAGE was computed using pretrained sex-specific models developed by applying support vector regression (SVR) with a radial kernel function and 10-fold cross-validation to 150 regional measures of cortical thickness (n = 68) and surface area (n = 68) and subcortical volume (n = 14), extracted with the Freesurfer Suite from a pooled sample of 18,965 healthy individuals (age range: 40-90 years; 54% females).^28^ Both the scripts and the models can be freely accessed at https://centilebrain.org/#/brainAge_global. These pre-trained models were applied to the brain morphometric data extracted from the UKB sample in this study to determine the G-brainAGE of each participant.

L-brainAGE estimates were computed using pre-trained voxel-based models developed by Popescu and colleagues^25^ from an independent sample of 4,155 healthy individuals (age range: 18-90 years). These pre-trained models were applied to the grey and white matter segments extracted from the brain scans of the current study sample.

### Non-imaging Measures

Initially, a total of 113 variables (Supplemental Table S1) were selected based on prior literature on brain aging^14,15^ pertaining to lifestyle (i.e., current diet, nutritional supplements, physical activity, alcohol consumption, smoking history, pastimes, use of electronics and screentime, sleep quality, quality of relationships) and to physical health and fitness (i.e., BMI, grip strength, etc.). Of these, only 55 variables were retained in the subsequent analyses; excluded variables had limited variability (i.e., >90% of the sample endorsed the same response) or high missingness (i.e., >10% missing values). In the retained 55 variables, missing values were imputed using Multivariate Imputation by Chained Equations implemented with the “mice” R package.^29^

### Statistical Approach

We used established procedures in HYDRA (https://github.com/evarol/HYDRA)^21^ to identify clusters of individuals with a history of depression, in reference to the non-psychiatric group. Factors with more than 2 levels were recoded to dummy variables, resulting in a total of 224 non-imaging measures as input features (Supplementary Table S2), and sex and age were modelled as covariates. HYDRA is a semi-supervised machine learning tool that clusters cases (here the depression group) based on their differences from a reference sample (here the non-psychiatric group) by finding multiple linear hyperplanes, which together form a convex polytope, thus extending linear max-margin classifiers to the non-linear space. We used 5-fold cross-validation to determine the clustering stability for a cluster range of 2-4, balancing model complexity while maintaining sufficient sample sizes in each cluster. The solution with the highest adjusted Rand index (ARI) was chosen. ARI is a statistical measure that quantifies similarity between data clusters and is adjusted for the chance grouping of datapoints. G-BrainAGE measures, socioeconomic status, and psychopathology were not used for clustering but to compare the resulting clusters using analysis of variance models (Supplemental Table S3). Group differences in L-brainAGE were tested using a general linear model implemented in SPM12 with age as covariate. Statistical significance was set at P_FWE_ < .05 after family-wise error (FWE) correction, and only clusters consisting of at least 400 voxels were considered.

### Involvement of Individuals with Lived Experience of Depression

This study was informed by focus groups comprising individuals with lived experience of depression, including users of the local mood disorder services. These focus groups identified the importance of understanding how lifestyle and fitness behaviours shape brain health in depression and their input directly influenced the study’s conceptual focus. This integration of experiential knowledge ensured that analyses using the UK Biobank resource were both contextually meaningful and clinically relevant.

## RESULTS

### HYDRA Detected Depression Clusters

HYDRA identified four clusters (ARI = 0.439; Supplemental Figure S2) within the depression group based on lifestyle factors and physical fitness. The cluster solution was similar regardless of modelling sex as a covariate (Supplementary Material, Section 5). The differentiation of the clusters was driven by 39 variables comprising social support and activities, physical exercise, fitness, smoking history, alcohol use, dietary patterns, nutritional supplements, computer and TV use, sleep quality, and outdoor exposure (Table 1). In addition, the clusters differed in mood-related symptoms, area deprivation, and education (Table 2) but not in chronological age or sex (Table 1).

**Table 1.** Cluster differences in the HYDRA input features among adults with a history of depression. Asterisks (*) indicate significant group effects that remained after correcting for multiple comparisons (P_FDR_ < .05). Variable types and test statistics are indicated by superscript letters.

|  | Cluster 1 | Cluster 2 | Cluster 3 | Cluster 4 | Statistic | df | P |
| --- | --- | --- | --- | --- | --- | --- | --- |
| **HYDRA COVARIATES** |  |  |  |  |  |  |  |
| Age<sup>a</sup> (years) | 63.08 | 63.08 | 63.50 | 62.31 | 0.790 | 3, 892 | .500 |
| Sex<sup>b</sup> (male) | 38.74% | 29.78% | 34.60% | 30.00% | 5.061 | 3 | .167 |
| **HYDRA INPUT FEATURES** |  |  |  |  |  |  |  |
| **Diet** |  |  |  |  |  |  |  |
| Beef intake<sup>c</sup> | 35.97% | 18.54% | 43.49% | 32.00% | 31.999 | 3 | <.001* |
| Lamb/mutton intake<sup>c</sup> | 18.58% | 3.93% | 20.00% | 10.67% | 28.251 | 3 | <.001* |
| Pork intake<sup>c</sup> | 19.76% | 10.11% | 31.75% | 18.00% | 34.469 | 3 | <.001* |
| Poultry intake<sup>c</sup> | 0.00% | 0.00% | 0.00% | 0.00% | - | - | - |
| Processed meat intake<sup>c</sup> | 29.64% | 9.55% | 31.11% | 24.67% | 31.519 | 3 | <.001* |
| Non-oily fish intake<sup>c</sup> | 16.60% | 8.99% | 11.75% | 16.67% | 7.344 | 3 | .062 |
| Oily fish intake<sup>c</sup> | 19.76% | 18.54% | 16.51% | 24.00% | 3.811 | 3 | .283 |
| Cheese intake<sup>c</sup> | 13.04% | 21.91% | 13.97% | 22.67% | 11.410 | 3 | .010* |
| Milk type mainly used<sup>d</sup> | 70.36% Semi-skimmed | 42.13% Semi-skimmed | 63.17% Semi-skimmed | 51.33% Semi-skimmed | 72.741 | 15 | <.001* |
| Spread type mainly used<sup>d</sup> | 60.47% Butter | 41.57% Butter | 55.87% Butter | 52.00% Butter | 17.887 | 6 | .007* |
| Dried fruit intake<sup>c</sup> | 56.52% | 56.74% | 29.52% | 44.67% | 53.953 | 3 | <.001* |
| Fresh fruit intake<sup>c</sup> | 37.55% | 43.26% | 23.17% | 36.00% | 25.008 | 3 | <.001* |
| Cooked vegetable intake<sup>c</sup> | 24.51% | 34.27% | 19.37% | 22.67% | 13.999 | 3 | .003* |
| Salad / raw vegetable intake<sup>c</sup> | 44.66% | 38.20% | 25.71% | 23.33% | 31.569 | 3 | <.001* |
| Bread intake<sup>c</sup> | 49.01% | 47.19% | 49.84% | 45.33% | 0.967 | 3 | .809 |
| Bread type mainly eaten<sup>d</sup> | 71.54% Wholegrain | 74.16% Wholegrain | 54.60% Wholegrain | 70.00% Wholegrain | 78.865 | 9 | <.001* |
| Cereal intake<sup>c</sup> | 51.78% | 47.75% | 35.24% | 46.00% | 17.258 | 3 | .001* |
| Cereal type mainly eaten<sup>d</sup> | 66.40% Muesli or oat (tied) | 35.39% Oat | 32.06% Don't eat cereal | 33.33% Oat | 86.202 | 15 | <.001* |
| Tea intake<sup>c</sup> | 51.38% | 41.01% | 43.17% | 46.67% | 5.736 | 3 | .125 |
| Water intake<sup>c</sup> | 41.11% | 60.67% | 35.56% | 36.00% | 33.098 | 3 | <.001* |
| Consume sugar<sup>b</sup> | 77.08% | 78.65% | 82.54% | 82.00% | 3.206 | 3 | .361 |
| Added salt intake<sup>c</sup> | 46.25% | 28.09% | 47.62% | 40.67% | 20.165 | 3 | <.001* |
| Major diet changes in past 5 years<sup>d</sup> | 64.82% No | 52.81% No | 49.52% No | 52.67% No | 29.882 | 6 | <.001* |
| Weekly variation in diet<sup>c</sup> | 5.93% | 8.99% | 11.11% | 14.00% | 8.086 | 3 | .044 |
| Nutritional supplements |  |  |  |  |  |  |  |
| Fish oil<sup>b</sup> | 22.53% | 28.09% | 20.32% | 19.33% | 4.924 | 3 | .177 |
| Glucosamine<sup>b</sup> | 12.25% | 18.54% | 5.08% | 12.00% | 22.280 | 3 | <.001* |
| Vitamin D<sup>b</sup> | 21.34% | 18.54% | 15.56% | 19.33% | 3.245 | 3 | .355 |
| Multivitamins +/- minerals<sup>b</sup> | 17.39% | 31.46% | 22.54% | 27.33% | 12.840 | 3 | .005* |
| Physical exercise |  |  |  |  |  |  |  |
| Heavy DIY (last 4 weeks)<sup>c</sup> | 37.94% | 42.13% | 37.14% | 27.33% | 8.122 | 3 | .044 |
| Light DIY (last 4 weeks)<sup>c</sup> | 48.62% | 33.71% | 31.11% | 36.00% | 20.081 | 3 | <.001* |
| Strenuous sports (last 4 weeks)<sup>c</sup> | 11.07% | 6.74% | 4.76% | 6.00% | 8.905 | 3 | .031* |
| Walking for pleasure (last 4 weeks)<sup>c</sup> | 49.01% | 46.07% | 31.11% | 44.67% | 22.045 | 3 | <.001* |
| Other exercise (last 4 weeks)<sup>c</sup> | 52.96% | 55.62% | 17.78% | 48.00% | 104.344 | 3 | <.001* |
| Weekly sports club / gym participation<sup>b</sup> | 46.25% | 29.78% | 13.33% | 32.00% | 74.772 | 3 | <.001* |
| Alcohol use and smoking |  |  |  |  |  |  |  |
| Alcohol intake frequency<sup>c</sup> | 52.17% | 39.89% | 43.81% | 18.67% | 45.033 | 3 | <.001* |
| Alcohol usually taken with meals<sup>d</sup> | 45.06% Yes | 44.38% Yes | 36.51% Yes | 60.67% Don't drink alcohol | 523.268 | 9 | <.001* |
| Smoking status<sup>d</sup> | 62.06% Never | 69.10% Never | 45.71% Never | 54.00% Never | 58.534 | 6 | <.001* |
| Social activities / relationships |  |  |  |  |  |  |  |
| Pub or social club<sup>b</sup> | 30.43% | 24.72% | 24.13% | 9.33% | 23.694 | 3 | <.001* |
| Religious group<sup>b</sup> | 17.00% | 14.61% | 10.48% | 26.67% | 20.436 | 3 | <.001* |
| Other weekly group activity<sup>b</sup> | 45.45% | 53.37% | 27.30% | 27.33% | 46.494 | 3 | <.001* |
| No regular social activities<sup>b</sup> | 13.44% | 24.16% | 43.81% | 26.67% | 66.345 | 3 | <.001* |
| Frequency of friend/ family visits<sup>c</sup> | 49.80% | 39.89% | 46.67% | 44.67% | 4.309 | 3 | .230 |
| Frequency of being able to confide<sup>c</sup> | 51.38% | 42.13% | 36.19% | 44.67% | 13.430 | 3 | .004* |
| Often feels lonely<sup>b</sup> | 13.04% | 37.08% | 36.51% | 29.33% | 45.733 | 3 | <.001* |
| Outdoor exposure |  |  |  |  |  |  |  |
| Time outdoors in the summer<sup>c</sup> | 45.85% | 56.74% | 41.59% | 40.00% | 12.856 | 3 | .005* |
| Time outdoors in the winter<sup>c</sup> | 47.83% | 56.74% | 37.46% | 41.33% | 18.785 | 3 | <.001* |
| Use of electronics / screentime |  |  |  |  |  |  |  |
| Length of mobile use<sup>c</sup> | 0.00% | 0.00% | 0.00% | 0.00% | - | - | - |
| Weekly mobile use<sup>c</sup> | 35.97% | 44.38% | 40.32% | 38.67% | 3.219 | 3 | .359 |
| Plays computer games<sup>b</sup> | 18.58% | 17.98% | 26.35% | 22.67% | 6.937 | 3 | .074 |
| Time spent using a computer<sup>c</sup> | 28.85% | 39.33% | 49.52% | 39.33% | 25.072 | 3 | <.001* |
| Time spent watching TV<sup>c</sup> | 33.20% | 29.78% | 43.49% | 35.33% | 11.394 | 3 | .010* |
| Sleep quality |  |  |  |  |  |  |  |
| Sleep duration<sup>a</sup> (hours) | 7.31 | 7.31 | 7.12 | 7.35 | 1.876 | 3, 892 | .132 |
| Insomnia<sup>c</sup> | 31.62% | 37.64% | 43.81% | 42.67% | 9.844 | 3 | .020* |
| Physical health |  |  |  |  |  |  |  |
| BMI<sup>a</sup> | 26.36 | 26.87 | 28.62 | 27.17 | 13.863 | 3, 892 | <.001* |
| Grip strength<sup>a</sup> | 28.18 | 26.48 | 27.25 | 25.26 | 3.025 | 3, 892 | .029* |
| <sup>a</sup>Continuous variable; values are means and *F* test statistic (one-way analysis of variance; ANOVA).<br><sup>b</sup>Dichotomous variable; values are percentage who endorse the measure and $\chi^2$ test statistic (test for relationship between cluster and response).<br><sup>c</sup>Ordinal variable; values are percentage above the pooled median and $\chi^2$ test statistic (test for relationship between cluster and percentage above the median).<br><sup>d</sup>Nominal variable; values are categories with the highest percentage of endorsement and $\chi^2$ test statistic (test for relationship between cluster and response distributions). Response distributions for nominal variables are provided in Supplemental Table S4 and Figure S3.<br>*$P_{FDR} < .05$ | | | | | | | |

**Table 2.**
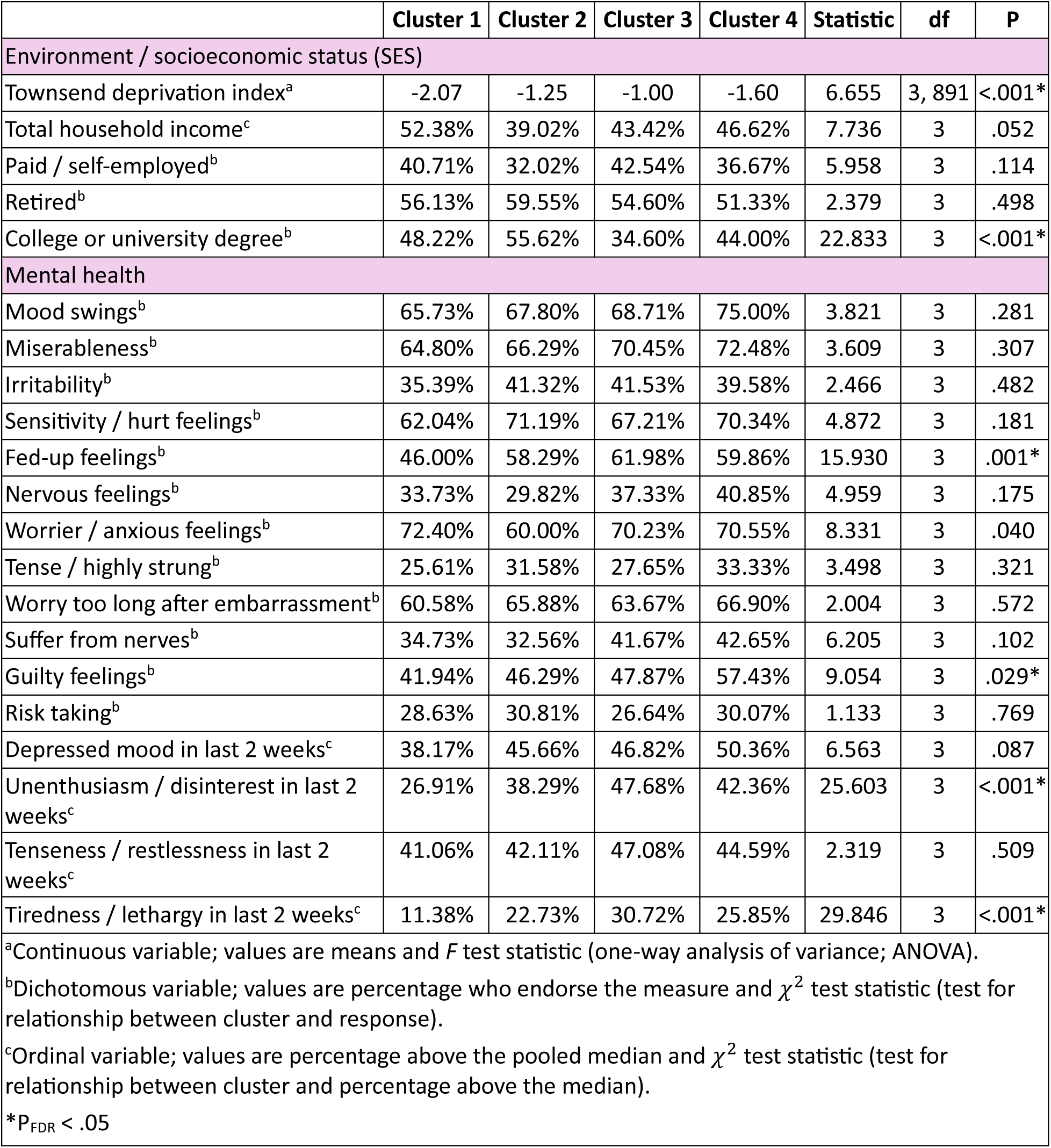
Cluster differences in socioeconomic indicators and psychopathology among adults with a history of major depression. Asterisks (*) indicate significant group effects that remained after correcting for multiple comparisons (P_FDR_ < .05). Variable types and test statistics are indicated by superscript letters.

#### Cluster 1

Cluster 1 (n = 253; 38.74% male) was the “balanced moderates” cluster, comprising individuals with strong social support, regular physical exercise, good physical fitness, the least amount of insomnia, low percentage of smokers, alcohol intake consumed with meals, a varied but stable diet, lowest computer use, and a moderate amount of outdoor exposure (Figure 1, purple bars; Supplemental Figure S3).

**Figure 1.**
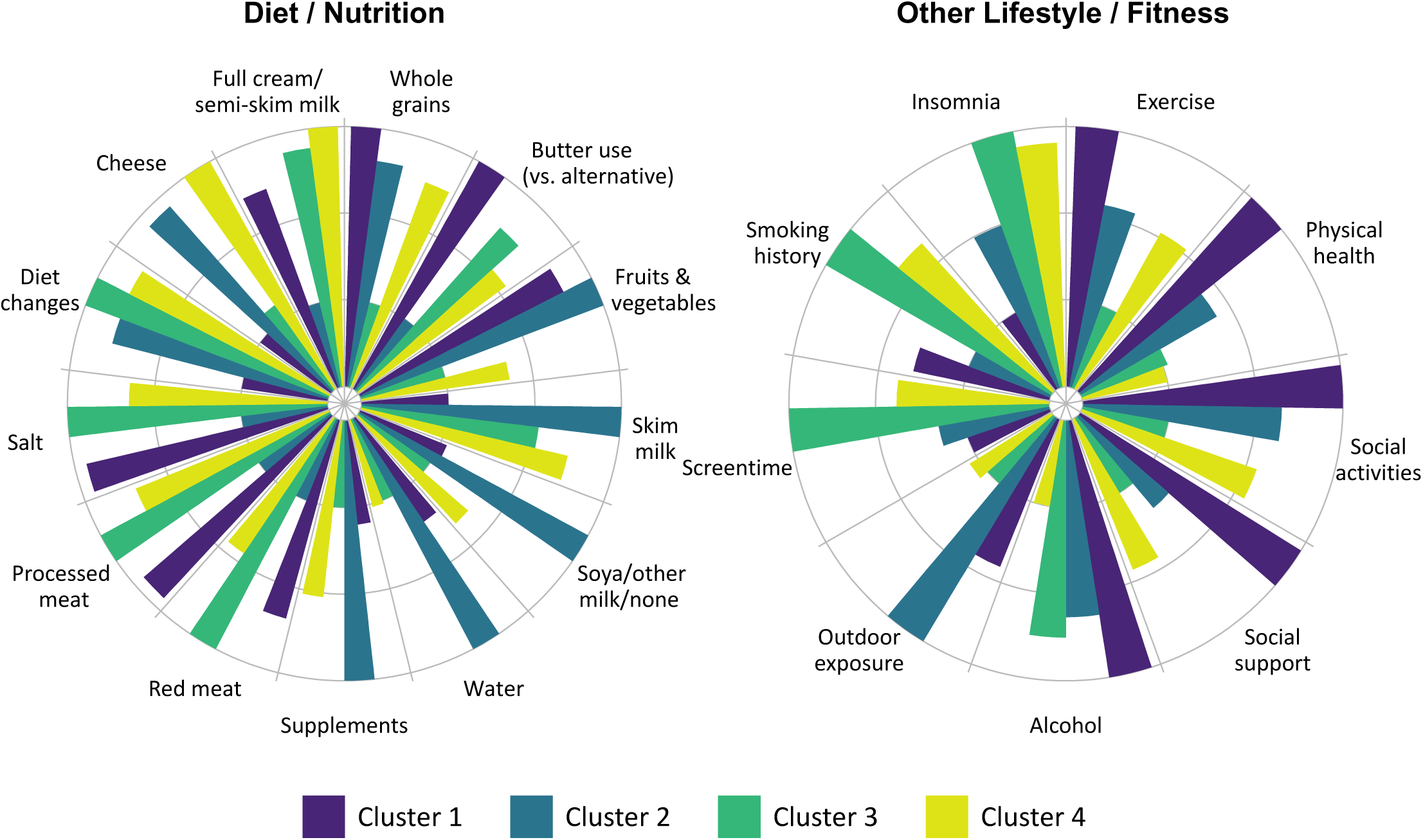
Visualization of HYDRA-based profiles of the clusters of individuals with a depression history. Values are group summary measures (e.g., mean) that have been scaled for ease of comparison to range from 1 (cluster with the lowest value) to 3 (cluster with the highest value) for variables with significant group differences (P_FDR_ < .05). Variables comprising composite measures include: (1) Whole grains (intake of whole grain bread, muesli cereal, and oat cereal), (2) Fruits & vegetables (intake of fresh fruit, dry fruit, salad/raw vegetables, and cooked vegetables), (3) Soya/other milk/none (preference for soya milk, other milk, or no milk), (4) Supplements (glucosamine and multivitamins), (5) Red meat (intake of pork, beef, and lamb), (6) Full cream/semi-skim milk (preference for full cream or semi-skimmed milk), (7) Exercise (gym participation, light DIY, walking for pleasure, strenuous and other exercise), (8) Physical health (low BMI and high grip strength), (9) Social support (low loneliness and high ability to confide in others), (10) Outdoor exposure (time spent outdoors in winter and in summer), and (11) Screentime (TV and computer use). Cluster 1 = “balanced moderates”; Cluster 2 = “optimal diet and activity”, Cluster 3 = “metabolic risk-sedentary”; Cluster 4 = “frailty-low activity”.

#### Cluster 2

Cluster 2 (n = 178; 29.78% male) was the “optimal diet and activity” cluster which comprised individuals with food choices consistent with healthy dietary guidelines^30^ emphasizing low meat and salt intake and high intake of fruit, vegetables, water, and dietary supplements (glucosamine and multivitamins). The cluster also had the lowest percentage of individuals with a smoking history (past or present), lowest TV use, and most outdoor exposure in both the summer and winter (Figure 1, blue bars; Supplemental Figure S3). In addition, individuals in this cluster were most likely to have a university education (Figure 2).

**Figure 2.**
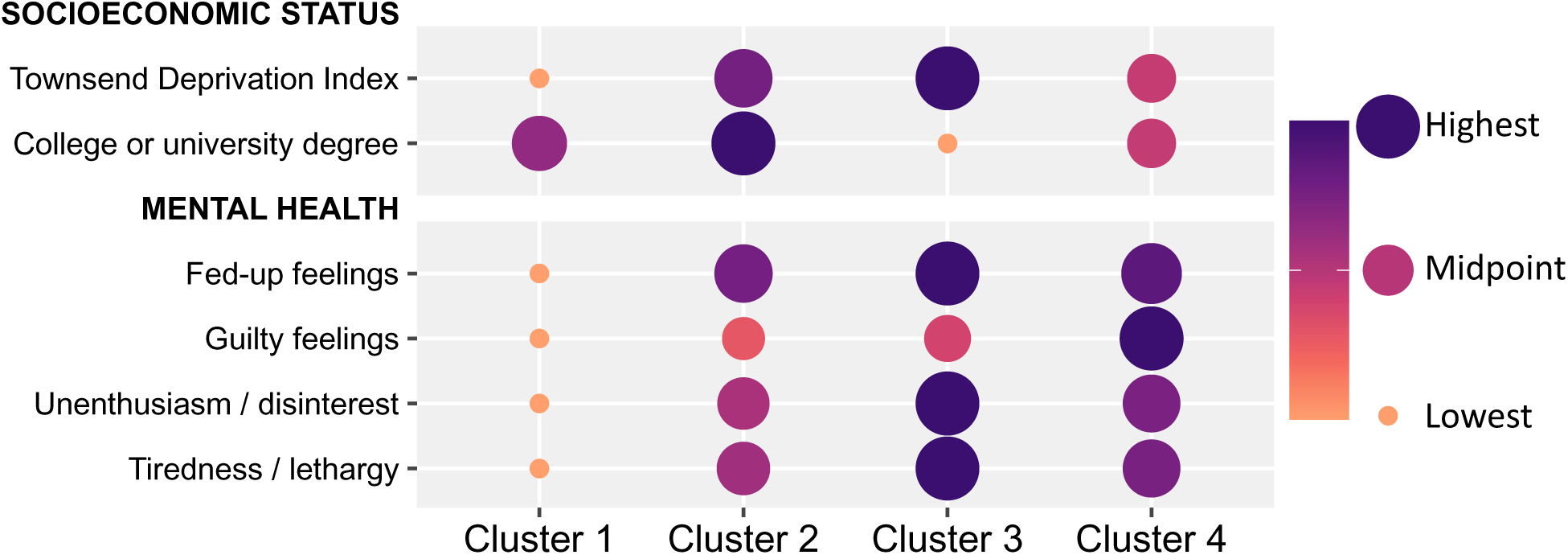
Profiles distinguishing depression clusters with respect to socioeconomic indicators and psychopathology ratings. Values are group summary measures (e.g., mean) that have been scaled for ease of comparison to range from 1 (cluster with the lowest value) to 3 (cluster with the highest value) for each variable with significant group differences (P_FDR_ < .05). Cluster 1 = “balanced moderates”; Cluster 2 = “optimal diet and activity”, Cluster 3 = “metabolic risk-sedentary”; Cluster 4 = “frailty-low activity”.

#### Cluster 3

Cluster 3 (n = 315; 34.60% male) was the “metabolic risk-sedentary” cluster which comprised individuals with diets based on high intake of red and processed meat, added salt, and low intake of fruit, vegetables, cereals, water, and dietary supplements. This cluster also had a mostly sedentary lifestyle characterized by low frequency of physical exercise and social activities, high amount of computer and TV use, and minimal outdoor exposure especially in the winter. The physical state of this cluster was also suboptimal and characterized by the highest amount of insomnia and highest BMI (Figure 1, green bars; Supplemental Figure S3). Additionally, individuals in Cluster 3 were more likely to live in socially deprived areas and had the lowest percentage of individuals with university education (Figure 2).

#### Cluster 4

Cluster 4 (n = 150; 30.00% male) was the “frailty-low activity” cluster which comprised individuals with low levels of alcohol use and a varied diet with a predominance of dairy products. Individuals in this cluster also had suboptimal physical health characterized by physical frailty inferred from their lower grip strength (Figure 1, yellow bars; Supplemental Figure S3).

### Cluster Differences in Mood Symptoms

Cluster differences in mood-related psychopathology are reported in Table 2 and Figure 2. The “balanced moderates” cluster (Cluster 1) had minimal psychopathology. The “optimal diet and activity” cluster (Cluster 2) reported mostly moderate fed-up feelings. The “metabolic risk-sedentary” (Cluster 3) and the “frailty-low activity” (Cluster 4) clusters were comparable and had the highest levels of mood-related psychopathology.

### Cluster Differences in G-brainAGE and L-BrainAGE

The mean G-brainAGE of the non-psychiatric group was near zero (−0.01), confirming that this group did not show evidence of deviation from the typical pattern of age-related brain changes. The age prediction model performance in the non-psychiatric group was comparable to that of prior replication samples^28^ with a mean absolute error (MAE) of 4.20 years, and a correlation between predicted and observed age of r = 0.72, P < .001. The G-brainAGE of the whole depression group was 0.31 years (approximately 4 calendar months) which was significantly higher compared to the non-psychiatric group (T(929.41) = 2.04, P = .042). The G-brainAGE of the four depression clusters did not differ from each other (all Ps > .05).

Compared to the non-psychiatric group, the whole depression group had significantly higher L- brainAGE in two clusters (P_FWE_ < .05; Figure 3). The largest cluster was centred in the anterior cingulate and extended to several medial and subcallosal frontal regions. The second cluster was restricted to the right insula. In terms of the specific depression clusters, only the “metabolic risk-sedentary” cluster (Cluster 3) had higher L-brainAGE compared to the non-psychiatric group in three voxel clusters respectively centred within the left parahippocampal gyrus, the right thalamus, and the left middle frontal gyrus, (P_FWE_ < .05; Figure 4).

**Figure 3.**
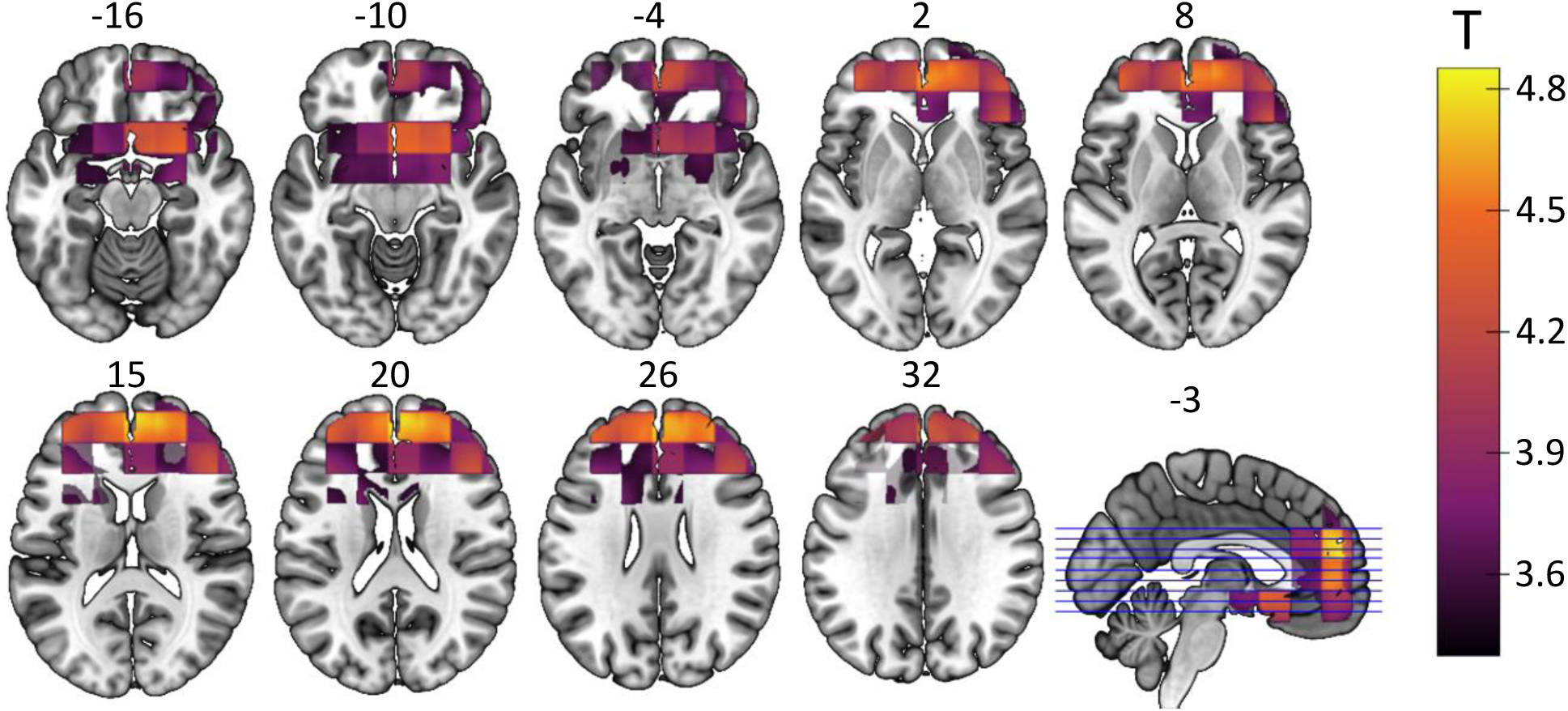
L-brainAGE differences between the non-psychiatric and the depression group. T-value overlay of statistically significant group differences in L-brainAGE (P_FWE_ < .05). Purple/yellow: depression group > non-psychiatric group. Images are displayed in neurological orientation with MNI coordinates.

**Figure 4.**
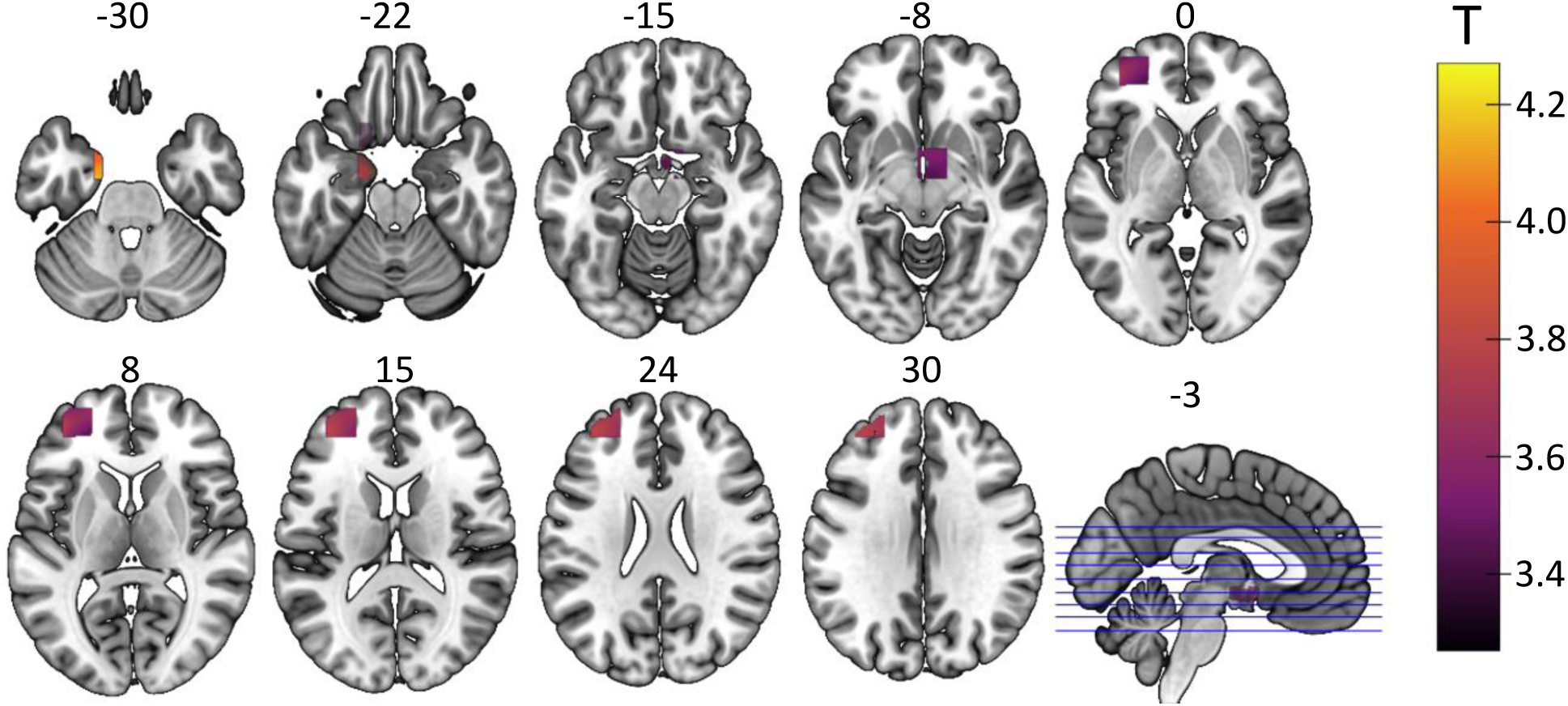
L-brainAGE differences between the non-psychiatric group and the “metabolic risk-sedentary” cluster. T-value overlay of statistically significant group differences in L-brainAGE (P_FWE_ < .05). Purple/yellow: “metabolic risk-sedentary” cluster 3 > non-psychiatric group. Images are displayed in neurological orientation with MNI coordinates.

## DISCUSSION

The present study stratified individuals with a history of the depression into four lifestyle and fitness based clusters: “balanced moderate,” “optimal diet and activity,” “metabolic risk-sedentary,” and “frail-low activity”. While all clusters exhibited higher G-brainAGE, the “metabolic risk-sedentary” cluster also showed older L-brainAGE in the frontotemporal regions. The clusters highlighted disparities in lifestyle, fitness, and socioeconomic status, with individuals in the “balanced moderates” and “optimal diet and activity” clusters benefiting from healthier living and lower psychopathology, linked to higher education and relatively prosperous living conditions. Conversely, the “metabolic risk-sedentary” and “frail-low activity” clusters, associated with lower socioeconomic status, demonstrated distinct patterns of unhealthy lifestyle patterns, cardiometabolic risks, and physical frailty, coupled with significant mood-related psychopathology. These findings emphasize that lifestyle and socioeconomic factors influence brain health in people with depression, in a fashion similar to that observed in the general population.

### Heterogeneity in Lifestyle and Fitness Factors Among Adults with a History of Depression

The study findings suggest that both positive and negative lifestyle choices and traits are not evenly distributed among individuals with a history of depression but tend to segregate into distinct clusters. The “balanced moderates” and the “optimal diet and activity” clusters included individuals that appeared to attend to general principles of healthy living, with the “balanced moderates” showing an advantage in terms of lower levels of psychopathology. Both clusters included individuals with higher levels of education living in more prosperous areas. These findings reinforce the association between socioeconomic status and healthier living which is not restricted to people with depression but is seen also in general population samples in the UK^31,32^ and elsewhere.^33^

A novel finding is that people with a history of depression and lower socioeconomic status are not a monolithic group, as we identified two different clusters within this population. The “metabolic risk-sedentary” cluster showed a constellation of cardiometabolic risk factors pertaining to poor diet and sedentary lifestyle which have previously been associated with lower socioeconomic circumstances.^34,35^ However, the “frail-low activity” cluster reported a varied diet but low physical activity which may be a consequence of suboptimal physical well-being. Both clusters, however, reported greater mood-related psychopathology which probably underscores the association between low socio-economic circumstances and depression.^36,37^

### Diet and Mood Symptoms Among Adults with a History of Depression

We noted four dietary patterns, concerning meat and fruit/vegetables, wholegrain foods, dairy products, and the use of nutritional supplements with distinct associations with mood symptoms. First, the “balanced moderates” cluster, which displayed the lowest levels of mood-related psychopathology, reported the highest consumption of salad and raw vegetables compared to the other clusters, aligning with existing evidence that a higher intake of raw fruits and vegetables is beneficial for mental health.^38^ However, findings from the “optimal diet and activity” cluster suggest a more nuanced picture. This cluster exhibited moderate levels of mood-related psychopathology, and reported minimal meat consumption, elevated intake of fruits and vegetables, and a relatively high preference for soy milk over full-fat or semi-skimmed milk, distinguishing it from the other three clusters. While direct evidence for nutritional deficiencies is unavailable in the present analyses, it is plausible that a connection exists between a low animal protein diet and depressive symptoms as indicated by another study conducted within the UKB which reported a higher prevalence of depression and lower dietary intakes of vitamins B6 and B12 among vegetarians compared to non-vegetarians.^39^ On the other extreme, the “metabolic risk-sedentary” cluster, which was characterized by the highest level of mood-related symptoms, reported high consumption of red meat and processed meats alongside reduced intake of fruits and vegetables, consistent with prior reports linking a high intake of red and processed meat to an increased risk of depression.^40^

Second, the “balanced moderates” cluster consumed more cereals than the other three clusters, particularly muesli and oats, as well as wholegrain brain. In contrast, the “metabolic risk - sedentary” cluster had the lowest intake of cereal and a marked preference for white bread over any other type of bread, setting it apart from the other three clusters. The physical health benefits of consuming whole grains are well-established^41^ and some evidence suggests that consumption of whole grain foods rather than refined grains may be associated with lower depression and anxiety, particularly in women.^42^ However, this association could also signify adherence to an overall healthier diet.

Third, the “balanced moderates” cluster, which exhibited the lowest levels of psychopathology, also reported the lowest cheese consumption and showed a preference for semi-skimmed milk over skimmed or full-cream milk. This preference was less pronounced in the other three clusters. The “frail-low activity” cluster, characterized by a moderate level of psychopathology, exhibited the highest cheese consumption and full cream, compared to the other three clusters. The consumption of low-fat dairy^43,44^ and fermented dairy foods like cheese and yogurt have been specifically linked to a reduced risk of depression.^45^ The finding in the “balanced moderates” cluster is consistent with these observations but the pattern observed in the “frail-low activity” cluster suggests that other traits and lifestyle choices present in this cluster may outweigh potential benefits.

Fourth, interpreting the use of nutritional supplements presents a particular challenge, as the underlying motivations remain unclear—it could reflect a need to address nutritional deficiencies or indicate heightened focus on overall health. Both explanations may account for the high levels of multivitamins/minerals supplement in the “optimal diet and activity” cluster. The lowest utilization was observed in the “balanced moderates” cluster, which may reflect their overall moderate approach to diet choices.

### Contextualizing Alcohol Use Among Adults with a History of Depression

The “balanced moderates” cluster exhibited the highest intake of alcohol with meals and in the context of social activities involving visits to pubs. This suggests that the alcohol intake within this cluster may be tied to social interactions rather than excessive alcohol use. It is also plausible that the cluster’s other health-promoting behaviours, including physical exercise and healthy dietary choices, serve as protective factors mitigating any potential negative impacts of alcohol intake. This underscores the intricate interplay of various dietary and lifestyle factors, highlighting how healthy practices can counterbalance less favourable ones.

### Older BrainAGE in Adults with a History of Depression

Compared to the non-psychiatric group, the depression group as a whole exhibited older G- brainAGE and higher L-brainAGE in frontal areas that was especially pronounced in medial and ventral prefrontal cortex including the subcallosal gyrus. These observations are meaningful given the relevance of these regions in the pathophysiology of depression due to their role in modulating emotional behaviour within cortico-limbic networks.^46^ The global as well as reward-related connectivity of the medial prefrontal cortex are blunted in individuals with depression^47,48^ while structural changes in these regions have been linked to the course of major depression.^49^ The older L-brainAGE of the subcallosal gyrus is also interesting as this region has been identified as both a site of dysfunction and a promising therapeutic target. Individuals with major depression exhibit reduced grey matter volume in this region,^50,51^ and functional connectivity within this area has been linked to varying treatment outcomes in depression.^52,53^ Longstanding recommendations advocate targeting the subcallosal gyrus in deep brain stimulation for depression,^54,55^ particularly in treatment-resistant cases.^56,57^

Notably, the different clusters were not associated with differential patterns of global and local brainAGE, except for the “metabolic risk-sedentary” cluster which exhibited older L-brainAGE compared to the non-psychiatric group not only in prefrontal areas but also in the parahippocampal gyrus and thalamus. This could reflect a particular vulnerability in these regions to lifestyle factors like poor diet and sedentary behaviour, as demonstrated in previous research highlighting the susceptibility of the hippocampal complex to metabolic risks and physical inactivity.^58–60^

### Limitations

The study is cross-sectional, and as such, it does not address either causality or the longitudinal evolution of the reported findings. The list of modifiable lifestyle variables and physical health measures considered was substantial but not exhaustive. The subset of participants with a known history of depression was smaller than the overall UKB sample, and although substantial, it may still have insufficient statistical power to fully disambiguate the heterogeneity of depression. Finally, to ensure broad applicability to older adults with a history of depression, age and sex were included as covariates to prevent clusters from reflecting lifestyle and physical fitness patterns primarily driven by these factors rather than depression history per se. This was particularly important for sex, given the higher occurrence of depression in females in both our sample and the general population.^61^

### Conclusions

This study advances a more nuanced understanding of the complex nature of depression by identifying distinct clusters of individuals with a history of the condition and examining their relationships with lifestyle, physical fitness factors, and age-related brain changes. The findings highlight the potential benefits of adopting a balanced, moderate lifestyle while underscoring the harmful effects of poor diet and low physical activity. Importantly, aside from the impact of poor diet and low physical activity, other lifestyle and fitness factors were not directly linked to older global or localized brainAGE. However, a history of depression—irrespective of cluster assignment—was associated with a slight deviation from typical age-related brain changes. Further research is needed to clarify the underlying mechanisms and develop targeted interventions to promote health in individuals with a history of depression.

## Data Availability

Access to the underlying data is available by application to the UK Biobank (https://www.ukbiobank.ac.uk/). The models for computing brainAGE from the neuroimaging data are freely available for both G-brainAGE (https://centilebrain.org/#/brainAge_global) and L-brainAGE (https://github.com/SebastianPopescu/U-NET-for-LocalBrainAge-prediction).

## Contributors

NS, YY, RG, and SSH contributed to data curation and methodology. NS performed all formal analyses and visualizations. NS and SF wrote the initial draft of the manuscript. LAMG and SF contributed to funding and resource acquisition. All authors contributed to reviewing and editing the manuscript. All authors had full access to all the data in the study and had final responsibility for the decision to submit for publication.

## Declaration of Interests

All authors declare no competing interests.

## Data Sharing

Access to the underlying data is available by application to the UK Biobank (https://www.ukbiobank.ac.uk/). The models for computing brainAGE from the neuroimaging data are freely available for both G-brainAGE (https://centilebrain.org/#/brainAge_global) and L- brainAGE (https://github.com/SebastianPopescu/U-NET-for-LocalBrainAge-prediction).

## Acknowledgements

Funding for this study was provided by the Canadian Institutes of Health Research and the Centre for Addiction and Mental Health *womenmind* foundation (both to LAMG). The work was supported by the computational resources provided by the Advanced Research Computing facilities at the University of British Columbia.

## Notes

### Competing Interest Statement

The authors have declared no competing interest.

### Funding Statement

This study was funded by the Canadian Institutes of Health Research (CIHR; Project Grant PJT186290) and the Centre for Addiction and Mental Health (CAMH) *womenmind* foundation.

### Author Declarations

Use of the study data was approved under UK Biobank data application number 69022. The UK Biobank has ethical approval from the Northwest Multi-Centre Research Ethics Committee (MREC). Further details on the UK Biobank Ethics and Governance framework are available here: https://www.ukbiobank.ac.uk/learn-more-about-uk-biobank/governance.

## REFERENCES

1 Setiawan E, Wilson AA, Mizrahi R, et al. Increased translocator protein distribution volume, a marker of neuroinflammation, in the brain during major depressive episodes. JAMA Psychiatry 2015; 72: 275.

2 Slavich GM. Psychoneuroimmunology of stress and mental health. In: Harkness K, Hayden EP, eds. The Oxford Handbook of Stress and Mental Health. New York: Oxford University Press, 2019. DOI:10.1093/oxfordhb/9780190681777.013.24.

3 Cole JH, Franke K. Predicting age using neuroimaging: Innovative brain ageing biomarkers. Trends Neurosci 2017; 40: 681–90.

4 Franke K, Gaser C. Ten years of BrainAGE as a neuroimaging biomarker of brain aging: What insights have we gained? Front Neurol 2019; 10: 789.

5 Han LKM, Schnack HG, Brouwer RM, et al. Contributing factors to advanced brain aging in depression and anxiety disorders. Transl Psychiatry 2021; 11: 402.

6 Han LKM, Dinga R, Hahn T, et al. Brain aging in major depressive disorder: Results from the ENIGMA major depressive disorder working group. Mol Psychiatry 2021; 26: 5124–39.

7 Jha MK, Chin Fatt C, Minhajuddin A, Mayes TL, Trivedi MH. Accelerated brain aging in adults with major depressive disorder predicts poorer outcome with sertraline: Findings from the EMBARC study. Biol Psychiatry Cogn Neurosci Neuroimaging 2023; 8: 462–70.

8 Ballester PL, Suh JS, Nogovitsyn N, et al. Accelerated brain aging in major depressive disorder and antidepressant treatment response: A CAN-BIND report. Neuroimage Clin 2021; 32: 102864.

9 Christman S, Bermudez C, Hao L, et al. Accelerated brain aging predicts impaired cognitive performance and greater disability in geriatric but not midlife adult depression. Transl Psychiatry 2020; 10: 317.

10 Ho NCW, Bethlehem RAI, Seidlitz J, et al. Atypical brain aging and its association with working memory performance in Major Depressive Disorder. Biol Psychiatry Cogn Neurosci Neuroimaging 2024; 9: 786–99.

11 Dunlop K, Victoria LW, Downar J, Gunning FM, Liston C. Accelerated brain aging predicts impulsivity and symptom severity in depression. Neuropsychopharmacology 2021; 46: 911–9.

12 Schiweck C, Valles-Colomer M, Arolt V, et al. Depression and suicidality: A link to premature T helper cell aging and increased Th17 cells. Brain Behav Immun 2020; 87: 603–9.

13 Simon MS, Ioannou M, Arteaga-Henríquez G, et al. Premature T cell aging in major depression: A double hit by the state of disease and cytomegalovirus infection. Brain Behav Immun Health 2023; 29: 100608.

14 Salih A, Boscolo Galazzo I, Raisi-Estabragh Z, et al. Brain age estimation at tract group level and its association with daily life measures, cardiac risk factors and genetic variants. Sci Rep 2021; 11: 20563.

15 Cole JH. Multimodality neuroimaging brain-age in UK biobank: Relationship to biomedical, lifestyle, and cognitive factors. Neurobiol Aging 2020; 92: 34–42.

16 Köhler CA, Evangelou E, Stubbs B, et al. Mapping risk factors for depression across the lifespan: An umbrella review of evidence from meta-analyses and Mendelian randomization studies. J Psychiatr Res 2018; 103: 189–207.

17 Schuch FB, Vancampfort D, Firth J, et al. Physical activity and incident depression: A meta-analysis of prospective cohort studies. Am J Psychiatry 2018; 175: 631–48.

18 Sarris J, Thomson R, Hargraves F, et al. Multiple lifestyle factors and depressed mood: A cross-sectional and longitudinal analysis of the UK Biobank (N = 84,860). BMC Med 2020; 18: 354.

19 Li Y, Lv MR, Wei YJ, et al. Dietary patterns and depression risk: A meta-analysis. Psychiatry Res 2017; 253: 373–82.

20 Sudlow C, Gallacher J, Allen N, et al. UK Biobank: An open access resource for identifying the causes of a wide range of complex diseases of middle and old age. PLoS Med 2015; 12: e1001779.

21 Varol E, Sotiras A, Davatzikos C. HYDRA: Revealing heterogeneity of imaging and genetic patterns through a multiple max-margin discriminative analysis framework. Neuroimage 2017; 145: 346–64.

22 Schuit AJ, Van Loon AJM, Tijhuis M, Ocké MC. Clustering of lifestyle risk factors in a general adult population. Prev Med (Baltim) 2002; 35: 219–24.

23 Morris LJ, D’Este C, Sargent-Cox K, Anstey KJ. Concurrent lifestyle risk factors: Clusters and determinants in an Australian sample. Prev Med (Baltim) 2016; 84: 1–5.

24 Watts P, Buck D, Netuveli G, Renton A. Clustering of lifestyle risk behaviours among residents of forty deprived neighbourhoods in London: Lessons for targeting public health interventions. J Public Health (Bangkok) 2016; 38: 308–15.

25 Popescu SG, Glocker B, Sharp DJ, Cole JH. Local brain-age: A U-Net model. Front Aging Neurosci 2021; 13: 761954.

26 Wang X, Hoffstaedter F, Kasper J, Eickhoff SB, Patil KR, Dukart J. Lifetime exposure to depression and neuroimaging measures of brain structure and function. JAMA Netw Open 2024; 7: e2356787.

27 Ashburner J. A fast diffeomorphic image registration algorithm. Neuroimage 2007; 38: 95–113.

28 Yu Y, Cui HQ, Haas SS, et al. Brain-age prediction: Systematic evaluation of site effects, and sample age range and size. Hum Brain Mapp 2024; 45: e26768.

29 van Buuren S, Groothuis-Oudshoorn K. mice: Multivariate Imputation by Chained Equations in R. J Stat Softw 2011; 45: 1–67.

30 U.S. Department of Agriculture and U.S. Department of Health and Human Services. Dietary Guidelines for Americans, 2020-2025, 9th edn. 2020 https://www.DietaryGuidelines.gov (accessed Oct 3, 2024).

31 Asamane EU; A;, Osei-Kwasi HA;, Boateng D, et al. Socioeconomic determinants of cardiovascular diseases, obesity, and diabetes among migrants in the United Kingdom: A systematic review. International Journal of Environmental Research and Public Health 2022, Vol 19, Page 3070 2022; 19: 3070.

32 Pampel FC, Krueger PM, Denney JT. Socioeconomic disparities in health behaviors. Annu Rev Sociol 2010; 36: 349–70.

33 Rippin HL, Hutchinson J, Greenwood DC, et al. Inequalities in education and national income are associated with poorer diet: Pooled analysis of individual participant data across 12 European countries. PLoS One 2020; 15: e0232447.

34 Burton NW, Turrell G, Oldenburg B. Participation in recreational physical activity: Why do socioeconomic groups differ? Health Education & Behavior 2003; 30: 225–44.

35 Nazri NS, Vanoh D, Leng SK. Malnutrition, low diet quality and its risk factors among older adults with low socio-economic status: A scoping review. Nutr Res Rev 2021; 34: 107–16.

36 Kok R, Avendano M, d’Uva TB, Mackenbach J. Can reporting heterogeneity explain differences in depressive symptoms across Europe? Soc Indic Res 2012; 105: 191–210.

37 Lorant V, Deliège D, Eaton W, Robert A, Philippot P, Ansseau M. Socioeconomic inequalities in depression: A meta-analysis. Am J Epidemiol 2003; 157: 98–112.

38 Brookie KL, Best GI, Conner TS. Intake of raw fruits and vegetables is associated with better mental health than intake of processed fruits and vegetables. Front Psychol 2018; 9: 487.

39 Berkins S, Schiöth HB, Rukh G. Depression and vegetarians: Association between dietary vitamin b6, b12 and folate intake and global and subcortical brain volumes. Nutrients 2021; 13: 1790.

40 Nucci D, Fatigoni C, Amerio A, Odone A, Gianfredi V. Red and processed meat consumption and risk of depression: A systematic review and meta-analysis. Int J Environ Res Public Health 2020; 17: 6686.

41 Miller KB. Review of whole grain and dietary fiber recommendations and intake levels in different countries. Nutr Rev 2020; 78: 29–36.

42 Sadeghi O, Hassanzadeh-Keshteli A, Afshar H, Esmaillzadeh A, Adibi P. The association of whole and refined grains consumption with psychological disorders among Iranian adults. Eur J Nutr 2019; 58: 211–25.

43 Cui Y, Huang C, Momma H, et al. Consumption of low-fat dairy, but not whole-fat dairy, is inversely associated with depressive symptoms in Japanese adults. Soc Psychiatry Psychiatr Epidemiol 2017; 52: 847–53.

44 Ghodsi R, Rostami H, Parastouei K, Taghdir M. Associations between whole and low-fat dairy products consumption, physical performance and mental health. Med J Nutrition Metab 2021; 14: 127–36.

45 Luo Y, Li Z, Gu L, Zhang K. Fermented dairy foods consumption and depressive symptoms: A meta-analysis of cohort studies. PLoS One 2023; 18: e0281346.

46 Rigucci S, Serafini G, Pompili M, Kotzalidis GD, Tatarelli R. Anatomical and functional correlates in major depressive disorder: The contribution of neuroimaging studies. The World Journal of Biological Psychiatry 2010; 11: 165–80.

47 Murrough JW, Abdallah CG, Anticevic A, et al. Reduced global functional connectivity of the medial prefrontal cortex in major depressive disorder. Hum Brain Mapp 2016; 37: 3214–23.

48 Rupprechter S, Romaniuk L, Series P, et al. Blunted medial prefrontal cortico-limbic reward-related effective connectivity and depression. Brain 2020; 143: 1946–56.

49 Dusi N, Barlati S, Vita A, Brambilla P. Brain structural effects of antidepressant treatment in major depression. Curr Neuropharmacol 2015; 13: 458–65.

50 Buchholz F, Meffert M, Bazin PL, Trampel R, Turner R, Schönknecht P. Highfield imaging of the subgenual anterior cingulate cortex in uni-and bipolar depression. Front Psychiatry 2024; 15: 1462919.

51 Yücel K, McKinnon MC, Chahal R, et al. Anterior cingulate volumes in never-treated patients with major depressive disorder. Neuropsychopharmacology 2008; 33: 3157–63.

52 Dunlop BW, Rajendra JK, Craighead WE, et al. Functional connectivity of the subcallosal cingulate cortex and differential outcomes to treatment with cognitive-behavioral therapy or antidepressant medication for major depressive disorder. American Journal of Psychiatry 2017; 174: 533–45.

53 Wang Y, Wang C, Zhou J, et al. Contribution of resting-state functional connectivity of the subgenual anterior cingulate to prediction of antidepressant efficacy in patients with major depressive disorder. Transl Psychiatry 2024; 14: 399.

54 Miller JP, Selman WR. Editorial: Deep brain stimulation for depression. J Neurosurg 2009; 111: 1207–8.

55 Teixeira SA, Moreira JL de S, Sousa NRT, et al. Molecular basis and clinical perspectives of deep brain stimulation for major depressive disorder. Journal of Cerebral Blood Flow & Metabolism 2022; 42: 683–5.

56 Khairuddin S, Ngo FY, Lim WL, et al. A decade of progress in deep brain stimulation of the subcallosal cingulate for the treatment of depression. Journal of Clinical Medicine 2020, Vol 9, Page 3260 2020; 9: 3260.

57 Puigdemont D, Pérez-Egea R, Portella MJ, et al. Deep brain stimulation of the subcallosal cingulate gyrus: Further evidence in treatment-resistant major depression. International Journal of Neuropsychopharmacology 2012; 15: 121–33.

58 Adachi Y, Ota K, Minami I, Yamada T, Watanabe T. Lower insulin secretion is associated with hippocampal and parahippocampal gyrus atrophy in elderly patients with type 2 diabetes mellitus. J Diabetes Investig 2021; 12: 1908–13.

59 Canário N, Crisóstomo J, Duarte JV, et al. Irreversible atrophy in memory brain regions over 7 years is predicted by glycemic control in type 2 diabetes without mild cognitive impairment. Front Aging Neurosci 2024; 16: 1367563.

60 Loprinzi PD. The effects of physical exercise on parahippocampal function. Physiol Int 2019; 106: 114–27.

61 Hoth KF, Voorhies K, Wu AC, Lange C, Potash JB, Lutz SM. The role of sex in genetic association studies of depression. J Psychiatr Brain Sci 2022; 7: e220013.

