## Supplemental for "Lifestyle and BrainAGE in Adult Depression"

Supplemental Material

### 1. Sample Selection

**Figure S1. Selection of participants included in the study.**

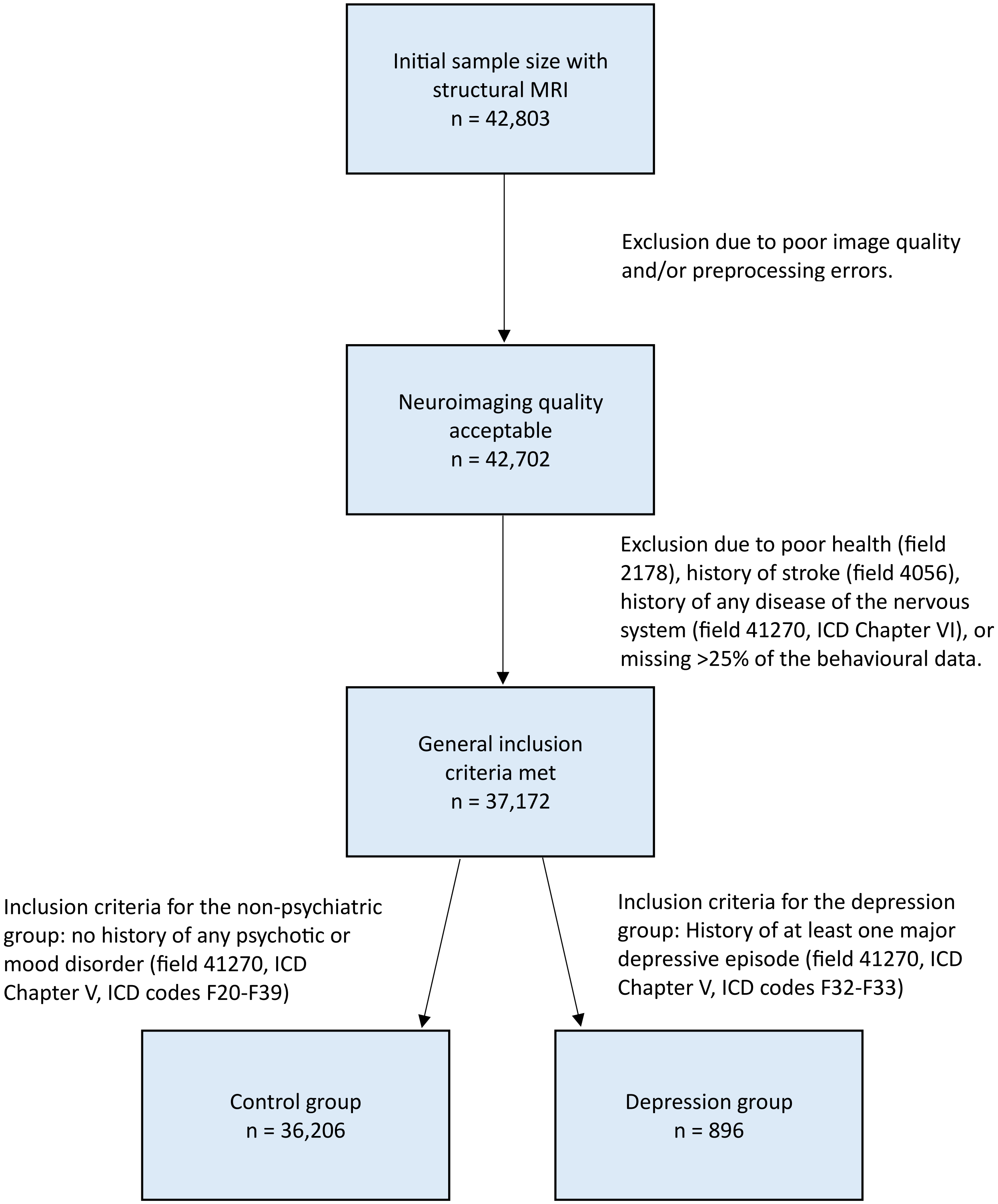

### 2. Selection of HYDRA Non-imaging Measures

#### Table S1. Non-imaging measures initially considered for inclusion as HYDRA features.

| Variable | Field ID | Category ID | Included in HYDRA | Reason for exclusion |
|---|---|---|---|---|
| Diet and nutrition | | | | |
| Beef intake | 1369 | 100052 (Diet) | yes | n/a |
| Lamb/mutton intake | 1379 | 100052 (Diet) | yes | n/a |
| Pork intake | 1389 | 100052 (Diet) | yes | n/a |
| Poultry intake | 1359 | 100052 (Diet) | yes | n/a |
| Processed meat intake | 1349 | 100052 (Diet) | yes | n/a |
| Non-oily fish intake | 1339 | 100052 (Diet) | yes | n/a |
| Oily fish intake | 1329 | 100052 (Diet) | yes | n/a |
| Cheese intake | 1408 | 100052 (Diet) | yes | n/a |
| Milk type used | 1418 | 100052 (Diet) | yes | n/a |
| Spread type | 1428 | 100052 (Diet) | yes | n/a |
| Dried fruit intake | 1319 | 100052 (Diet) | yes | n/a |
| Fresh fruit intake | 1309 | 100052 (Diet) | yes | n/a |
| Cooked vegetable intake | 1289 | 100052 (Diet) | yes | n/a |
| Salad / raw vegetable intake | 1299 | 100052 (Diet) | yes | n/a |
| Bread intake | 1438 | 100052 (Diet) | yes | n/a |
| Bread type | 1448 | 100052 (Diet) | yes | n/a |
| Cereal intake | 1458 | 100052 (Diet) | yes | n/a |
| Cereal type | 1468 | 100052 (Diet) | yes | n/a |
| Tea intake | 1488 | 100052 (Diet) | yes | n/a |
| Water intake | 1528 | 100052 (Diet) | yes | n/a |
| Never eat eggs, dairy, wheat, sugar (multiple selections allowed) | 6144 | 100052 (Diet) | - | - |
| – Sugar or foods/drinks containing sugar | 6144 | 100052 (Diet) | yes | n/a |
| – Eggs or food containing eggs | 6144 | 100052 (Diet) | no | 97.99% eat eggs or food containing eggs |
| – Dairy products | 6144 | 100052 (Diet) | no | 97.99% eat dairy products |
| – Wheat products | 6144 | 100052 (Diet) | no | 97.11% eat wheat products |
| – Eat all of the above | 6144 | 100052 (Diet) | no | Uninformative with 3/4 of the categories removed from analysis |
| Salt added to food | 1478 | 100052 (Diet) | yes | n/a |
| Major dietary changes in the last 5 years | 1538 | 100052 (Diet) | yes | n/a |

| Variable | Field ID | Category ID | Included in HYDRA | Reason for exclusion |
|---|---|---|---|---|
| Variation in diet | 1548 | 100052 (Diet) | yes | n/a |
| Mineral and other dietary supplements (multiple selections allowed) | 6179 | 100045 (Medication) | - | - |
| – Fish oil (including cod liver oil) | 6179 | 100045 (Medication) | yes | n/a |
| – Glucosamine | 6179 | 100045 (Medication) | yes | n/a |
| – Calcium | 6179 | 100045 (Medication) | no | 93.33% do not take calcium |
| – Zinc | 6179 | 100045 (Medication) | no | 95.80% do not take zinc |
| – Iron | 6179 | 100045 (Medication) | no | 96.61% do not take iron |
| – Selenium | 6179 | 100045 (Medication) | no | 98.22% do not take selenium |
| – None of the above | 6179 | 100045 (Medication) | no | Uninformative with 4/6 of the categories removed from analysis |
| Vitamin and mineral supplements (multiple selections allowed) | 6155 | 100045 (Medication) | - | - |
| – Vitamin D | 6155 | 100045 (Medication) | yes | n/a |
| – Multivitamins +/- minerals | 6155 | 100045 (Medication) | yes | n/a |
| – Vitamin A | 6155 | 100045 (Medication) | no | 98.59% do not take vitamin A |
| – Vitamin B | 6155 | 100045 (Medication) | no | 95.15% do not take vitamin B |
| – Vitamin C | 6155 | 100045 (Medication) | no | 92.70% do not take vitamin C |
| – Vitamin E | 6155 | 100045 (Medication) | no | 98.19% do not take vitamin E |
| – Folic acid or Folate (Vit B9) | 6155 | 100045 (Medication) | no | 98.18% do not take folic acid or folate |
| – None of the above | 6155 | 100045 (Medication) | no | Uninformative with 5/7 of the categories removed from analysis |
| Coffee intake | 1498 | 100052 (Diet) | no | Data unavailable |
| Coffee type | 1508 | 100052 (Diet) | no | Coffee intake data unavailable |
| Non-butter spread type details | 2654 | 100052 (Diet) | no | Low endorsement of individual types, and the consolidation of them into one "non-butter" category is already captured by "Spread type" (field 1428) |
| Physical activity | | | | |
| Frequency of heavy DIY in last 4 weeks | 2624 | 100054 (Physical activity) | yes | n/a |

| Variable | Field ID | Category ID | Included in HYDRA | Reason for exclusion |
|---|---|---|---|---|
| Frequency of light DIY in last 4 weeks | 1011 | 100054 (Physical activity) | yes | n/a |
| Frequency of strenuous sports in last 4 weeks | 991 | 100054 (Physical activity) | yes | n/a |
| Frequency of walking for pleasure in last 4 weeks | 971 | 100054 (Physical activity) | yes | n/a |
| Frequency of other exercises in last 4 weeks | 3637 | 100054 (Physical activity) | yes | n/a |
| Types of physical activity in last 4 weeks (multiple selections allowed) | 6164 | 100054 (Physical activity) | no | This variable was used to identify the types of activity that participants had engaged in in the last 4 weeks (see activities measured above), but it does not capture the amount of any given activity |
| Duration of heavy DIY | 2634 | 100054 (Physical activity) | no | Captured physical activity by frequency rather than duration |
| Duration of light DIY | 1021 | 100054 (Physical activity) | no | Captured physical activity by frequency rather than duration |
| Duration of moderate activity | 894 | 100054 (Physical activity) | no | Captured physical activity by frequency rather than duration |
| Duration of other exercises | 3647 | 100054 (Physical activity) | no | Captured physical activity by frequency rather than duration |
| Duration of strenuous sports | 1001 | 100054 (Physical activity) | no | Captured physical activity by frequency rather than duration |
| Duration of vigorous activity | 914 | 100054 (Physical activity) | no | Captured physical activity by frequency rather than duration |
| Duration of walks | 874 | 100054 (Physical activity) | no | Captured physical activity by frequency rather than duration |
| Duration walking for pleasure | 981 | 100054 (Physical activity) | no | Captured physical activity by frequency rather than duration |
| Number of days/week of moderate physical activity 10+ minutes | 884 | 100054 (Physical activity) | no | Captured by frequency of various physical activities in last 4 weeks |
| Number of days/week of vigorous physical activity 10+ minutes | 904 | 100054 (Physical activity) | no | Captured by frequency of various physical activities in last 4 weeks |
| Number of days/week walked 10+ minutes | 864 | 100054 (Physical activity) | no | Captured by frequency of various physical activities in last 4 weeks |
| Alcohol intake | | | | |
| Alcohol intake frequency | 1558 | 100051 (Alcohol) | yes | n/a |
| Alcohol usually taken with meals | 1618 | 100051 (Alcohol) | yes | n/a |

| Variable | Field ID | Category ID | Included in HYDRA | Reason for exclusion |
|---|---|---|---|---|
| Alcohol drinker status | 20117 | 100051 (Alcohol) | no | Alcohol intake is captured by "Alcohol intake frequency", field 1558 |
| Average monthly beer plus cider intake | 4429 | 100051 (Alcohol) | no | 78.79% missing |
| Average monthly champagne plus white wine intake | 4418 | 100051 (Alcohol) | no | 78.83% missing |
| Average monthly fortified wine intake | 4451 | 100051 (Alcohol) | no | 78.77% missing |
| Average monthly intake of other alcoholic drinks | 4462 | 100051 (Alcohol) | no | 78.86% missing |
| Average monthly red wine intake | 4407 | 100051 (Alcohol) | no | 78.84% missing |
| Average monthly spirits intake | 4440 | 100051 (Alcohol) | no | 78.79% missing |
| Average weekly beer plus cider intake | 1588 | 100051 (Alcohol) | no | 27.48% missing |
| Average weekly champagne plus white wine intake | 1578 | 100051 (Alcohol) | no | 27.59% missing |
| Average weekly fortified wine intake | 1608 | 100051 (Alcohol) | no | 27.51% missing |
| Average weekly intake of other alcoholic drinks | 5364 | 100051 (Alcohol) | no | 27.44% missing |
| Average weekly red wine intake | 1568 | 100051 (Alcohol) | no | 27.57% missing |
| Average weekly spirits intake | 1598 | 100051 (Alcohol) | no | 27.60% missing |
| Smoking | | | | |
| Smoking status | 20116 | 100058 (Smoking) | yes | n/a |
| Current tobacco smoking | 1239 | 100058 (Smoking) | no | 96.72% responded "no" |
| Number of cigarettes currently smoked daily (current cigarette smokers) | 3456 | 100058 (Smoking) | no | 98.30% missing or not applicable |
| Type of tobacco currently smoked | 3446 | 100058 (Smoking) | no | 96.72% do not currently smoke tobacco (field 1239) |
| Pastimes / leisure activities | | | | |
| Leisure/social activities (multiple selections allowed) | 6160 | 100061 (Social support) | - | - |
| – Sports club or gym | 6160 | 100061 (Social support) | yes | n/a |
| – Pub or social club | 6160 | 100061 (Social support) | yes | n/a |

| Variable | Field ID | Category ID | Included in HYDRA | Reason for exclusion |
|---|---|---|---|---|
| – Religious group | 6160 | 100061 (Social support) | yes | n/a |
| – Other group activity | 6160 | 100061 (Social support) | yes | n/a |
| – None of the above | 6160 | 100061 (Social support) | yes | n/a |
| – Adult education class | 6160 | 100061 (Social support) | (yes) | Combined with "Other group activity" due to low endorsement (92.28% responded "no") |
| Time spent outdoors in summer | 1050 | 100055 (Sun exposure) | yes | n/a |
| Time spent outdoors in winter | 1060 | 100055 (Sun exposure) | yes | n/a |
| Electronics / screentime | | | | |
| Length of mobile phone use | 1110 | 100053 (Electronic device use) | yes | n/a |
| Weekly usage of mobile phone in last 3 months | 1120 | 100053 (Electronic device use) | yes | n/a |
| Plays computer games | 2237 | 100053 (Electronic device use) | yes | n/a |
| Time spent using computer | 1080 | 100054 (Physical activity) | yes | n/a |
| Time spent watching television (TV) | 1070 | 100054 (Physical activity) | yes | n/a |
| Sleep quality | | | | |
| Sleep duration | 1160 | 100057 (Sleep) | yes | n/a |
| Sleeplessness / insomnia | 1200 | 100057 (Sleep) | yes | n/a |
| Daytime dozing / sleeping | 1220 | 100057 (Sleep) | no | Sleep quality already captured by sleep duration (field 1160) and amount of sleeplessness/insomnia (field 1200) |
| Getting up in morning | 1170 | 100057 (Sleep) | no | Sleep quality already captured by sleep duration (field 1160) and amount of sleeplessness/insomnia (field 1200) |
| Morning/evening person (chronotype) | 1180 | 100057 (Sleep) | no | Sleep quality already captured by sleep duration (field 1160) and amount of sleeplessness/insomnia (field 1200) |
| Nap during day | 1190 | 100057 (Sleep) | no | Sleep quality already captured by sleep duration (field 1160) and amount of sleeplessness/insomnia (field 1200) |
| Snoring | 1210 | 100057 (Sleep) | no | Sleep quality already captured by sleep duration (field 1160) and amount of sleeplessness/insomnia (field 1200) |
| Relationships / social support | | | | |
| Frequency of friend/family visits | 1031 | 100061 (Social support) | yes | n/a |
| Able to confide | 2110 | 100061 (Social support) | yes | n/a |

| Variable | Field ID | Category ID | Included in HYDRA | Reason for exclusion |
|---|---|---|---|---|
| Loneliness, isolation | 2020 | 100060 (Mental health) | yes | n/a |
| Physical measures | | | | |
| Body mass index (BMI) | 21001 | 100010 (Body size measures) | yes | n/a |
| Body mass index (BMI) | 23104 | 100009 (Body composition by impedance) | (yes) | Only used for a small subsample of participants (n=31) who were missing BMI data for body size measurements (field 21001) |
| Hand grip strength (left) | 46 | 100019 (Hand grip strength) | (yes) | Left and right hand measurements were averaged to compute one grip strength variable |
| Hand grip strength (right) | 47 | 100019 (Hand grip strength) | (yes) | Left and right hand measurements were averaged to compute one grip strength variable |
| Diastolic blood pressure, automated reading | 4079 | 100011 (Blood pressure) | no | 11.92% missing blood pressure data |
| Diastolic blood pressure, manual reading | 94 | 100011 (Blood pressure) | no | 11.92% missing blood pressure data |
| Systolic blood pressure, automated reading | 4080 | 100011 (Blood pressure) | no | 11.92% missing blood pressure data |
| Systolic blood pressure, manual reading | 93 | 100011 (Blood pressure) | no | 11.92% missing blood pressure data |

**Table S2. HYDRA variables (covariates and features).**

| Variable in UKB dataset | HYDRA feature | Field ID | Variable type |
|---|---|---|---|
| COVARIATES | | | |
| Age when attended assessment centre | Age | 21003 | continuous |
| Sex (female/male) | Sex (female/male) | 31 | dichotomous |
| FEATURES | | | |
| Diet | | | |
| Servings of beef per week | | 1369 | ordinal |
| | Never | | |
| | Less than once a week | | |
| | Once a week | | |
| | More than once a week (pooled responses for "2-4 times a week", "5-6 times a week", and "once or more daily") | | |
| Lamb/mutton intake (how often, not including processed meat) | | 1379 | ordinal |
| | Never | | |
| | Less than once a week | | |
| | At least once a week (pooled responses for "once a week", "2-4 times a week", "5-6 times a week", and "once or more daily") | | |
| Pork intake (how often, NOT including processed meats such as bacon or ham) | | 1389 | ordinal |
| | Never | | |
| | Less than once a week | | |
| | At least once a week (pooled responses for "once a week", "2-4 times a week", "5-6 times a week", and "once or more daily") | | |
| Poultry intake (how often, NOT including processed meats) | | 1359 | ordinal |
| | Never | | |
| | Less than once a week | | |
| | Once a week | | |
| | More than once a week (pooled responses for "2-4 times a week", "5-6 times a week", and "once or more daily") | | |

| Variable in UKB dataset | HYDRA feature | Field ID | Variable type |
|---|---|---|---|
| Processed meat intake (how often, e.g., bacon, ham, sausages, meat pies, kebabs, burgers, chicken nuggets) | | 1349 | ordinal |
| | Never | | |
| | Less than once a week | | |
| | Once a week | | |
| | More than once a week (pooled responses for "2-4 times a week", "5-6 times a week", and "once or more daily") | | |
| Non-oily fish intake (how often, e.g., cod, tinned tuna, haddock) | | 1339 | ordinal |
| | Never | | |
| | Less than once a week | | |
| | Once a week | | |
| | More than once a week (pooled responses for "2-4 times a week", "5-6 times a week", and "once or more daily") | | |
| Oily fish intake (how often, e.g., sardines, salmon, mackerel, herring) | | 1329 | ordinal |
| | Never | | |
| | Less than once a week | | |
| | Once a week | | |
| | More than once a week (pooled responses for "2-4 times a week", "5-6 times a week", and "once or more daily") | | |
| Cheese intake (how often, including cheese in pizzas, quiches, cheese sauce, etc.) | | 1408 | ordinal |
| | Never or less than once a week (pooled responses for "never" and "less than once a week") | | |
| | Once a week | | |
| | 2-4 times a week | | |
| | 5-6 times a week | | |
| | Once or more daily | | |
| Milk type mainly used | | 1418 | nominal |
| | Full cream | | |
| | Semi-skimmed | | |
| | Skimmed | | |
| | Soya | | |
| | Other type of milk | | |
| | Never/rarely have milk | | |
| Spread type (mainly used) | | 1428 | nominal |
| | Butter/spreadable butter | | |
| | Other type of spread/margarine | | |
| | Never/rarely use spread | | |

| Variable in UKB dataset | HYDRA feature | Field ID | Variable type |
|---|---|---|---|
| Dried fruit intake (pieces per day, e.g., 1 prune, 1 apricot, or 10 raisins would each count as one piece) | | 1319 | ordinal |
| | 0 | | |
| | < 1 | | |
| | 1 | | |
| | 2 | | |
| | 3 | | |
| | > 3 | | |
| Fresh fruit intake (pieces per day, e.g., 1 apple, 1 banana, or 10 grapes would each count as one piece) | | 1309 | ordinal |
| | 0 | | |
| | < 1 | | |
| | 1 | | |
| | 2 | | |
| | 3 | | |
| | 4 | | |
| | 5 | | |
| | > 5 | | |
| Cooked vegetable intake (heaped tablespoons per day) | | 1289 | ordinal |
| | 0 or < 1 | | |
| | 1 | | |
| | 2 | | |
| | 3 | | |
| | 4 | | |
| | 5 | | |
| | > 5 | | |
| Salad / raw vegetable intake (heaped tablespoons per day, including lettuce or tomato in sandwiches) | | 1299 | ordinal |
| | 0 | | |
| | < 1 | | |
| | 1 | | |
| | 2 | | |
| | 3 | | |
| | 4 | | |
| | 5 | | |
| | > 5 | | |
| Bread intake (slices per week) | | 1438 | ordinal |
| | 0 to <1 | | |
| | 1-4 | | |
| | 5-8 | | |
| | 9-12 | | |
| | 13-16 | | |
| | 17-20 | | |
| | >20 | | |

| Variable in UKB dataset | HYDRA feature | Field ID | Variable type |
|---|---|---|---|
| Bread type mainly eaten | | 1448 | nominal |
| | White | | |
| | Brown | | |
| | Wholemeal or wholegrain | | |
| | Other type of bread | | |
| Cereal intake (bowls per week) | | 1458 | ordinal |
| | 0 | | |
| | < 1 | | |
| | 1 | | |
| | 2 | | |
| | 3 | | |
| | 4 | | |
| | 5 | | |
| | 6 | | |
| | > 6 | | |
| Cereal type mainly eaten | | 1468 | nominal |
| | Biscuit cereal (e.g., Weetabix) | | |
| | Bran cereal (e.g., All Bran, Branflakes) | | |
| | Muesli | | |
| | Oat cereal (e.g., Ready Brek, porridge) | | |
| | Other (e.g., Cornflakes, Frosties) | | |
| | N/A, don't eat cereal (indicated by "Cereal intake", field 1458-2.0) | | |
| Tea intake (cups per day) | | 1488 | ordinal |
| | 0 or < 1 | | |
| | 1 | | |
| | 2 | | |
| | 3 | | |
| | 4 | | |
| | 5 | | |
| | 6 | | |
| | > 6 | | |

| Variable in UKB dataset | HYDRA feature | Field ID | Variable type |
|---|---|---|---|
| Water intake (glasses per day) | | 1528 | ordinal |
| | 0 | | |
| | < 1 | | |
| | 1 | | |
| | 2 | | |
| | 3 | | |
| | 4 | | |
| | 5 | | |
| | 6 | | |
| | > 6 | | |
| Never eat eggs, dairy, wheat, sugar (NEVER eat which of the following: eggs or foods containing eggs, dairy products, wheat products, sugar or foods/drinks containing sugar, or none (eat all of the above); multiple answers could be selected) | | 6144 | n/a, multiple selections allowed |
| | Sugar or foods/drinks containing sugar (yes/no) | | dichotomous |
| Salt added to food (not including salt used in cooking) | | 1478 | ordinal |
| | Never/rarely | | |
| | Sometimes | | |
| | Usually or always (pooled responses for "usually" and "always") | | |
| Major dietary changes in the last 5 years (any) | | 1538 | nominal |
| | No | | |
| | Yes, because of illness | | |
| | Yes, because of other reasons | | |
| Variation in diet (does it vary much from week to week) | | 1548 | ordinal |
| | Never/rarely | | |
| | Sometimes | | |
| | Often | | |
| Nutritional supplements | | | |
| Mineral and other dietary supplements (regularly take any of the following: fish oil, glucosamine, calcium, zinc, iron, selenium, or none; multiple answers could be selected) | | 6179 | n/a, multiple selections allowed |
| | Fish oil (yes/no) | | dichotomous |
| | Glucosamine (yes/no) | | dichotomous |
| Vitamin and mineral supplements (regularly take any of the following: vitamin A/B/C/D/E, folic acid / folcate (vit B9), multivitamins +/- minerals, or none; multiple answers could be selected) | | 6155 | n/a, multiple selections allowed |
| | Vitamin D (yes/no) | | dichotomous |
| | Multivitamins +/- minerals (yes/no) | | dichotomous |

| Variable in UKB dataset | HYDRA feature | Field ID | Variable type |
|---|---|---|---|
| Physical activity | | | |
| Frequency of heavy DIY in last 4 weeks | | 2624 | ordinal |
| | Never (derived from "Types of physical activity in last 4 weeks, field 6164) | | |
| | Once in the last 4 weeks | | |
| | 2-3 times in the last 4 weeks | | |
| | Once a week | | |
| | At least twice a week (pooled responses from "2-3 times a week", "4-5 times a week", and "every day") | | |
| Frequency of light DIY in last 4 weeks | | 1011 | ordinal |
| | Never or only once ("never" derived from "Types of physical activitiy in last 4 weeks, field 6164) | | |
| | 2-3 times in the last 4 weeks | | |
| | Once a week | | |
| | 2-3 times a week | | |
| | At least 4 times a week (pooled respones from "4-5 times a week" and "every day") | | |
| Frequency of strenuous sports in last 4 weeks | | 991 | ordinal |
| | Never (derived from "Types of physical activitiy in last 4 weeks, field 6164) | | |
| | Once a week or less (pooled responses from "once in the last 4 weeks", "2-3 times in the last 4 weeks", and "once a week") | | |
| | At least twice a week (pooled responses from "2-3 times a week", "4-5 times a week", and "every day") | | |
| Frequency of walking for pleasure in last 4 weeks | | 971 | ordinal |
| | Never or only once ("never" derived from "Types of physical activitiy in last 4 weeks, field 6164) | | |
| | 2-3 times in the last 4 weeks | | |
| | Once a week | | |
| | 2-3 times a week | | |
| | 4-5 times a week | | |
| | Every day | | |

| Variable in UKB dataset | HYDRA feature | Field ID | Variable type |
|---|---|---|---|
| Frequency of other exercises in last 4 weeks (e.g., swimming, cycling, keeping fit) | | 3637 | ordinal |
| | Never or only once ("never" derived from "Types of physical activitiy in last 4 weeks, field 6164) | | |
| | 2-3 times in the last 4 weeks | | |
| | Once a week | | |
| | 2-3 times a week | | |
| | At least 4 times a week (pooled respones from "4-5 times a week" and "every day") | | |
| Alcohol intake | | | |
| Alcohol intake frequency | | 1558 | ordinal |
| | Never | | |
| | Special occasions only | | |
| | One to three times a month | | |
| | Once or twice a week | | |
| | Three or four times a week | | |
| | Daily or almost daily | | |
| Alcohol usually taken with meals | | 1618 | nominal |
| | N/A, don't drink (indicated by response to "Alcohol intake frequency", field 1558) | | |
| | No | | |
| | It varies | | |
| | Yes | | |
| Smoking | | | |
| Smoking status | | 20116 | nominal |
| | Current | | |
| | Never | | |
| | Previous | | |
| Pastimes / leisure activities | | | |
| Leisure/social activities (attend which of the following at least once a week: sports club or gym, pub or social club, religious group, adult education class, other group activity, or none; multiple answers could be selected) | | 6160 | n/a, multiple selections allowed |
| | Pub or social club (yes/no) | | dichotomous |
| | Religious group (yes/no) | | dichotomous |
| | Sports club or gym (yes/no) | | dichotomous |
| | Other group activity (pooled with "adult education class") (yes/no) | | dichotomous |
| | None (yes/no) | | dichotomous |

| Variable in UKB dataset | HYDRA feature | Field ID | Variable type |
|---|---|---|---|
| Time spent outdoors in summer (hours/day) | | 1050 | ordinal |
| | 0-1 | | |
| | 2 | | |
| | 3 | | |
| | 4 | | |
| | 5 | | |
| | 6 | | |
| | > 6 | | |
| Time spent outdoors in winter (hours/day) | | 1060 | ordinal |
| | 0 to <1 | | |
| | 1 | | |
| | 2 | | |
| | 3 | | |
| | 4 | | |
| | > 4 | | |
| Electronics / screentime | | | |
| Length of mobile phone use (how many years using a mobile phone at least once per week to make or receive calls) | | 1110 | ordinal |
| | Never used mobile phone at least once per week | | |
| | No more than 4 years (pooled responses from "one year or less" and "two to four years") | | |
| | 5-8 years | | |
| | More than 8 years | | |
| Weekly usage of mobile phone in last 3 months | | 1120 | ordinal |
| | None | | |
| | Less than 5 minutes | | |
| | 5-29 minutes | | |
| | 30-59 minutes | | |
| | 1-3 hours | | |
| | > 3 hours (pooled responses from "4-6 hours" and "more than 6 hours") | | |
| Plays computer games | Plays computer games at least sometimes (yes/no; pooled responses for "sometimes" and "often") | 2237 | dichotomous |

| Variable in UKB dataset | HYDRA feature | Field ID | Variable type |
|---|---|---|---|
| Time spent using computer (hours per day) | | 1080 | ordinal |
| | 0 | | |
| | < 1 | | |
| | 1 | | |
| | 2 | | |
| | 3 | | |
| | > 3 | | |
| Time spent watching TV (hours per day) | | 1070 | ordinal |
| | 0 to <1 | | |
| | 1 | | |
| | 2 | | |
| | 3 | | |
| | 4 | | |
| | 5 | | |
| | > 5 | | |
| Sleep quality | | | |
| Sleep duration (approximate number of hours in every 24-hour period) | Sleep duration | 1160 | continuous |
| Sleeplessness / insomnia (frequency) | | 1200 | ordinal |
| | Never/rarely | | |
| | Sometimes | | |
| | Usually | | |
| Relationships / social support | | | |
| Frequency of friend/family visits | | 1031 | ordinal |
| | Never/almost never or no friends/family outside household (pooled responses for "never or almost never" and "no friends/family outside household) | | |
| | Once every few months | | |
| | About once a month | | |
| | About once a week | | |
| | 2-4 times a week | | |
| | Almost daily | | |
| Able to confide (frequency) | | 2110 | ordinal |
| | Never or almost never | | |
| | Once every few months | | |
| | About once a month | | |
| | About once a week | | |
| | 2-4 times a week | | |
| | Almost daily | | |
| Loneliness, isolation (often feels lonely) | Loneliness, isolation (yes/no) | 2020 | dichotomous |

| **Variable in UKB dataset** | **HYDRA feature** | **Field ID** | **Variable type** |
|---|---|---|---|
| Physical health | | | |
| Body mass index (BMI; kg/m$^2$); constructed from height and weight measurements where available (n = 35,910) OR impedance measurement (n = 31) | BMI | 21001 (height & weight measurement) or 23104 (impedance measurement) | continuous |
| Hand grip strength (kg force units) | Grip strength (averaged across both hands) | 46 (left grip) & 47 (right grip) | continuous |

### Table S3. Variables examined but not used for HYDRA Clustering

| Variable | Definition | Field ID | Variable type |
|---|---|---|---|
| BrainAGE | | | |
| G-brainAGE | Difference in years between a participant’s chronological age and their "brain age" predicted from their neuroimaging data, calculated using pretrained, sex-specific CentileBrain models. | n/a | continuous |
| Environment / socioeconomic status (SES) | | | |
| Townsend deprivation index at recruitment | Townsend deprivation index calculated immediately prior to participant joining UK Biobank. Based on the preceding national census output areas. Each participant is assigned a score corresponding to the output area in which their postcode is located. | 189 | continuous |
| Average total household income before tax | Options: "< £18K", "£18K-£30,999", "£31K-£51,999", "£52K-£100K", ">£100K" | 738 | ordinal |
| Current employment status (multiple selections allowed)[a] | | 6142 | n/a, multiple selections allowed |
| – In paid employment or self-employed | Yes/no | 6142 | dichotomous |
| – Retired | Yes/no | 6142 | dichotomous |
| College or university degree | Yes/no, extracted from UKB field "Qualifications" | 6138 | dichotomous |
| Mental health | | | |
| Mood swings | Yes/no, "Does your mood often go up and down?" | 1920 | dichotomous |
| Miserableness | Yes/no, "Do you ever feel 'just miserable' for now reason?" | 1930 | dichotomous |
| Irritability | Yes/no, "Are you an irritable person?" | 1940 | dichotomous |
| Sensitivity / hurt feelings | Yes/no, "Are your feelings easily hurt?" | 1950 | dichotomous |
| Fed-up feelings | Yes/no, "Do you often feel 'fed-up'?" | 1960 | dichotomous |
| Nervous feelings | Yes/no, "Would you call yourself a nervous person?" | 1970 | dichotomous |
| Worrier / anxious feelings | Yes/no, "Are you a worrier?" | 1980 | dichotomous |
| Tense / 'highly strung' | Yes/no, "Would you call yourself tense or 'highly strung'?" | 1990 | dichotomous |
| Worry too long after embarrassment | Yes/no, "Do you worry too long after an embarrassing experience?" | 2000 | dichotomous |
| Suffer from "nerves" | Yes/no, "Do you suffer from 'nerves'?" | 2010 | dichotomous |
| Guilty feelings | Yes/no, "Are you often troubled by feelings of guilt?" | 2030 | dichotomous |
| Risk taking | Yes/no, "Would you describe yourself as someone who takes risks?" | 2040 | dichotomous |

| Variable | Definition | Field ID | Variable type |
| --- | --- | --- | --- |
| Frequency of depressed mood in last 2 weeks | Options: "Not at all", "Several days", "More than half the days", "Nearly every day" | 2050 | ordinal |
| Frequency of unenthusiasm / disinterest in last 2 weeks | Options: "Not at all", "Several days", "More than half the days", "Nearly every day" | 2060 | ordinal |
| Frequency of tenseness / restlessness in last 2 weeks | Options: "Not at all", "Several days", "More than half the days", "Nearly every day" | 2070 | ordinal |
| Frequency of tiredness / lethargy in last 2 weeks | Options: "Not at all", "Several days", "More than half the days", "Nearly every day" | 2080 | ordinal |
| [a] Three items within this variable, “Looking after home and/or family”, “Unemployed”, “Full or part-time student”, “Unable to work because of sickness or disability”, and “Doing unpaid or voluntary work”, were removed from the analysis due to low variance in responses (each was < 10% endorsed). | | | |

#### 3. HYDRA Hyperparameter Tuning

Heterogeneity Through Discriminative Analysis (HYDRA) is a non-linear machine learning algorithm for integrated binary classification and subpopulation clustering using a multiple max-margin discriminative analysis framework (Varol et al., 2017). The detailed code can be found on https://github.com/evarol/HYDRA. Classification in HYDRA is based on indices of deviation between a clinical and the healthy reference group; healthy individuals are separated from the clinical sample using a convex polytope formed by combining multiple linear hyperplanes. The multiple hyperplanes model potential heterogeneity within clinical samples while their combination extends linear max-margin classifiers to the non-linear space.

We applied the HYDRA algorithm to identify subtypes within the depression group, using 5-fold cross-validation to ensure the stability of clustering solutions, indexed by the adjusted rand index (ARI) for cross-fold reproducibility. For hyperparameter selection, we largely adhered to the developers' default settings, as the algorithm showed good convergence. However, to optimize clustering stability, we tested a range of configurations for the number of clusters (k = 2, 3, 4), the regularization parameter (0.25, 0.50, 0.75, 1.0; promoting sparsity in the estimated hyperplanes), and initialization strategies, which included k-means assignment, pure random assignment, random hyperplanes, and determinantal point process (DPP) random hyperplanes. The best results, yielding the highest ARI values, were obtained with k-means initialization and a 4-cluster model (Figure S2). Adjusting the regularization parameter did not influence clustering stability (ARI values) across any values of k or initialization strategy.

**Figure S2. Adjusted Rand Indices (ARIs) for each value of k and initialization strategy.**

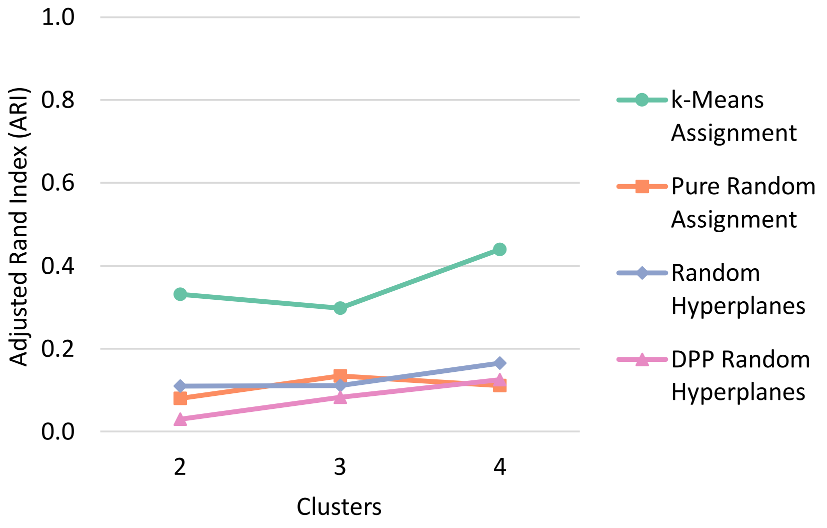

### 4. Additional HYDRA Results

**Table S4. Group descriptives for all variables.**

| | Control group | Whole depression group | Cluster 1 | Cluster 2 | Cluster 3 | Cluster 4 |
|---|---|---|---|---|---|---|
| HYDRA COVARIATES | | | | | | |
| Age (mean years) | 63.84 | 63.10 | 63.08 | 63.08 | 63.50 | 62.31 |
| Sex (% male) | 47.59% | 34.04% | 38.74% | 29.78% | 34.60% | 30.00% |
| HYDRA INPUT FEATURES | | | | | | |
| Diet | | | | | | |
| Beef intake (median servings/week) | < 1/week | < 1/week | < 1/week | None | < 1/week | < 1/week |
| Lamb/mutton intake (median servings/week) | < 1/week | < 1/week | < 1/week | None | < 1/week | < 1/week |
| Pork intake (median servings/week) | < 1/week | < 1/week | < 1/week | None | < 1/week | < 1/week |
| Poultry intake (median servings/week) | >1/week | >1/week | >1/week | 1/week | >1/week | >1/week |
| Processed meat intake (median servings/week) | 1/week | 1/week | 1/week | None | 1/week | 1/week |
| Non-oily fish intake (median servings/week) | 1/week | 1/week | 1/week | 1/week | 1/week | < 1/week |
| Oily fish intake (median servings/week) | 1/week | 1/week | 1/week | 1/week | < 1/week | 1/week |
| Cheese intake (median servings/week) | 2-4/week | 2-4/week | 2-4/week | 2-4/week | 2-4/week | 2-4/week |
| Milk type mainly used (% endorsement) | | | | | | |
| Full cream | 7.24% | 6.36% | 3.95% | 5.06% | 6.98% | 10.67% |
| Semi-skimmed | 63.51% | 59.04% | 70.36% | 42.13% | 63.17% | 51.33% |
| Skimmed | 19.22% | 21.65% | 16.21% | 26.97% | 21.90% | 24.00% |
| Soya | 3.18% | 4.24% | 4.35% | 8.43% | 1.90% | 4.00% |
| Other | 3.25% | 4.24% | 3.95% | 10.11% | 1.27% | 4.00% |
| Never/rarely drink milk | 3.61% | 4.46% | 1.19% | 7.30% | 4.76% | 6.00% |
| Spread type mainly used (% endorsement) | | | | | | |
| Butter | 56.00% | 53.68% | 60.47% | 41.57% | 55.87% | 52.00% |
| Other spread/ margarine | 33.22% | 34.93% | 31.23% | 41.57% | 33.33% | 36.67% |
| Never/rarely use spread | 10.79% | 11.38% | 8.30% | 16.85% | 10.79% | 11.33% |

| | Control group | Whole depression group | Cluster 1 | Cluster 2 | Cluster 3 | Cluster 4 |
|---|---|---|---|---|---|---|
| Dried fruit intake (median pieces/day) | < 1/day | none | < 1/day | < 1/day | none | None |
| Fresh fruit intake (median pieces/day) | 2/day | 2/day | 2/day | 2/day | 2/day | 2/day |
| Cooked vegetable intake (median tablespoons/day) | 3/day | 3/day | 3/day | 3/day | 2/day | 2/day |
| Salad / raw vegetable intake (median tablespoons/day) | 2/day | 2/day | 2/day | 2/day | 1/day | 1/day |
| Bread intake (median slices/week) | 5-8/week | 5-8/week | 5-8/week | 5-8/week | 5-8/week | 5-8/week |
| Bread type mainly eaten (% endorsement) | | | | | | |
| Wholemeal/wholegrain | 68.95% | 65.85% | 71.54% | 74.16% | 54.60% | 70.00% |
| Brown | 10.87% | 10.27% | 12.65% | 14.61% | 6.35% | 9.33% |
| White | 14.27% | 16.63% | 10.28% | 4.49% | 30.16% | 13.33% |
| Other | 5.91% | 7.25% | 5.53% | 6.74% | 8.89% | 7.33% |
| Cereal intake (median bowls/week) | 5/week | 5/week | 6/week | 5/week | 4/week | 5/week |
| Cereal type mainly eaten (% endorsement) | | | | | | |
| Bran | 10.79% | 8.59% | 9.49% | 6.18% | 9.21% | 8.67% |
| Oat | 28.82% | 29.46% | 33.20% | 35.39% | 21.27% | 33.33% |
| Muesli | 21.63% | 20.76% | 33.20% | 16.85% | 12.70% | 21.33% |
| Biscuit | 11.21% | 9.71% | 6.32% | 11.80% | 12.38% | 7.33% |
| Other | 8.25% | 9.04% | 6.72% | 5.62% | 12.38% | 10.00% |
| None | 19.30% | 22.43% | 11.07% | 24.16% | 32.06% | 19.33% |
| Tea intake (median cups/day) | 3/day | 3/day | 4/day | 3/day | 3/day | 3/day |
| Water intake (median glasses/day) | 2/day | 2/day | 2/day | 3/day | 2/day | 2/day |
| Consume sugar (% endorsement) | 79.61% | 80.13% | 77.08% | 78.65% | 82.54% | 82.00% |
| Added salt intake (median frequency) | Never/rarely | Never/rarely | Never/rarely | Never/rarely | Never/rarely | Never/rarely |
| Major diet changes in the past 5 years (% endorsement) | | | | | | |
| None | 64.51% | 55.02% | 64.82% | 52.81% | 49.52% | 52.67% |

| | Control group | Whole depression group | Cluster 1 | Cluster 2 | Cluster 3 | Cluster 4 |
|---|---|---|---|---|---|---|
| Yes due to illness | 5.18% | 9.60% | 3.16% | 10.11% | 15.24% | 8.00% |
| Yes for other reason | 30.31% | 35.38% | 32.02% | 37.08% | 35.24% | 39.33% |
| Weekly variation in diet (median frequency) | Sometimes | Sometimes | Sometimes | Sometimes | Sometimes | Sometimes |
| Nutritional supplements | | | | | | |
| Fish oil (% usage) | 25.13% | 22.32% | 22.53% | 28.09% | 20.32% | 19.33% |
| Glucosamine (% usage) | 13.09% | 10.94% | 12.25% | 18.54% | 5.08% | 12.00% |
| Vitamin D (% usage) | 15.39% | 18.42% | 21.34% | 18.54% | 15.56% | 19.33% |
| Multivitamins +/- minerals (% usage) | 19.91% | 23.66% | 17.39% | 31.46% | 22.54% | 27.33% |
| Physical exercise | | | | | | |
| Heavy DIY (median frequency, last 4 weeks) | Never | Never | Never | Never | Never | Never |
| Light DIY (median frequency, last 4 weeks) | 2-3x total | 2-3x total | 2-3x total | Never/only once | Never/only once | 2-3x total |
| Strenuous sports (median frequency, last 4 weeks) | Never | Never | Never | Never | Never | Never |
| Walking for pleasure (median frequency, last 4 weeks) | Once/week | Once/week | Once/week | Once/week | 2-3x total | Once/week |
| Other exercise (median frequency, last 4 weeks) | 2-3x total | Never/only once | 2-3x total | 2-3x total | Never/only once | Never/only once |
| Weekly sports club / gym participation (% participation) | 39.05% | 29.02% | 46.25% | 29.78% | 13.33% | 32.00% |
| Alcohol use and smoking | | | | | | |
| Alcohol intake frequency (median) | 1-2x/week | 1-2x/week | 3-4x/week | 1-2x/week | 1-2x/week | Never |
| Alcohol usually taken with meals (% endorsement) | | | | | | |
| Yes | 46.17% | 38.62% | 45.06% | 44.38% | 36.51% | 25.33% |
| No | 16.21% | 19.64% | 17.79% | 23.03% | 27.94% | 1.33% |
| Varies | 31.62% | 31.58% | 37.15% | 32.58% | 35.56% | 12.67% |
| Don't drink | 5.99% | 10.16% | 0.00% | 0.00% | 0.00% | 60.67% |
| Smoking status (% endorsement) | | | | | | |
| Current | 3.18% | 7.03% | 1.19% | 1.12% | 12.70% | 12.00% |
| Previous | 33.27% | 36.61% | 36.76% | 29.78% | 41.59% | 34.00% |

| | Control group | Whole depression group | Cluster 1 | Cluster 2 | Cluster 3 | Cluster 4 |
|---|---|---|---|---|---|---|
| Never | 63.55% | 56.36% | 62.06% | 69.10% | 45.71% | 54.00% |
| Social activities / relationships | | | | | | |
| Pub or social club (% participation) | 39.05% | 29.02% | 30.43% | 24.72% | 24.13% | 9.33% |
| Religious group (% participation) | 15.65% | 15.85% | 17.00% | 14.61% | 10.48% | 26.67% |
| Other weekly group activity (% participation) | 36.87% | 37.61% | 45.45% | 53.37% | 27.30% | 27.33% |
| No regular social activities (% endorsement) | 22.73% | 28.46% | 13.44% | 24.16% | 43.81% | 26.67% |
| Frequency of friend/ family visits (median) | Once/week | Once/week | Once/week | Once/week | Once/week | Once/week |
| Frequency of being able to confide (median) | Daily | 2-4x/week | Daily | 2-4x/week | Once/week | 2-4x/week |
| Often feels lonely (% endorsement) | 11.90% | 28.79% | 13.04% | 37.08% | 36.51% | 29.33% |
| Outdoor exposure | | | | | | |
| Time outdoors in the summer (median) | 3 hours/day | 3 hours/day | 3 hours/day | 4 hours/day | 3 hours/day | 3 hours/day |
| Time outdoors in the winter (median) | 1 hour/day | 1 hour/day | 1 hour/day | 2 hours/day | 1 hour/day | 1 hour/day |
| Use of electronics / screentime | | | | | | |
| Length of mobile use (median) | 9+ yrs | 9+ yrs | 9+ yrs | 9+ yrs | 9+ yrs | 9+ yrs |
| Weekly mobile use (median) | 5-29 min | 5-29 min | 5-29 min | 5-29 min | 5-29 min | 5-29 min |
| Plays computer games (% endorsement) | 19.05% | 21.88% | 18.58% | 17.98% | 26.35% | 22.67% |
| Time spent using a computer (median) | 1 hour/day | 1 hour/day | 1 hour/day | 1 hour/day | 1 hour/day | 1 hour/day |
| Time spent watching TV (median) | 3 hours/day | 3 hours/day | 3 hours/day | 2 hours/day | 3 hours/day | 3 hours/day |
| Sleep quality | | | | | | |
| Sleep duration (mean hours) | 7.16 | 7.25 | 7.31 | 7.31 | 7.12 | 7.35 |
| Insomnia (median frequency) | Sometimes | Sometimes | Sometimes | Sometimes | Sometimes | Sometimes |
| Physical health | | | | | | |
| BMI (mean $kg/m^2$) | 26.24 | 27.39 | 26.36 | 26.87 | 28.62 | 27.17 |

| | Control group | Whole depression group | Cluster 1 | Cluster 2 | Cluster 3 | Cluster 4 |
|---|---|---|---|---|---|---|
| Grip strength (mean kg force units) | 30.19 | 27.03 | 28.18 | 26.48 | 27.25 | 25.26 |
| VARIABLES NOT USED FOR HYDRA CLUSTERING | | | | | | |
| BrainAGE | | | | | | |
| G-brainAGE (mean years) | -0.01 | 0.31 | 0.30 | 0.38 | 0.14 | 0.65 |
| Environment / socioeconomic status | | | | | | |
| Townsend deprivation index (mean) | -1.93 | -1.45 | -2.07 | -1.25 | -1.00 | -1.60 |
| Total household income before tax (median) | £31,000 - £51,999 | £18,000 - £30,999 | £31,000 - £51,999 | £18,000 - £30,999 | £18,000 - £30,999 | £18,000 - £30,999 |
| Paid / self-employed (% endorsement) | 39.85% | 38.95% | 40.71% | 32.02% | 42.54% | 36.67% |
| Retired (% endorsement) | 59.12% | 55.47% | 56.13% | 59.55% | 54.60% | 51.33% |
| College or university degree (% endorsement) | 50.2% | 44.2% | 48.22% | 55.62% | 34.60% | 44.00% |
| Mental health | | | | | | |
| Mood swings (% endorsement) | 33.28% | 68.74% | 65.73% | 67.80% | 68.71% | 75.00% |
| Miserableness (% endorsement) | 31.99% | 68.37% | 64.80% | 66.29% | 70.45% | 72.48% |
| Irritability (% endorsement) | 24.11% | 39.42% | 35.39% | 41.32% | 41.53% | 39.58% |
| Sensitivity / hurt feelings (% endorsement) | 44.70% | 67.09% | 62.04% | 71.19% | 67.21% | 70.34% |
| Fed-up feelings (% endorsement) | 28.52% | 56.38% | 46.00% | 58.29% | 61.98% | 59.86% |
| Nervous feelings (% endorsement) | 17.12% | 35.38% | 33.73% | 29.82% | 37.33% | 40.85% |
| Worrier / anxious feelings (% endorsement) | 48.74% | 68.86% | 72.40% | 60.00% | 70.23% | 70.55% |
| Tense / highly strung (% endorsement) | 10.99% | 28.81% | 25.61% | 31.58% | 27.65% | 33.33% |
| Worry too long after embarrassment (% endorsement) | 46.67% | 63.79% | 60.58% | 65.88% | 63.67% | 66.90% |
| Suffer from nerves (% endorsement) | 13.54% | 38.02% | 34.73% | 32.56% | 41.67% | 42.65% |
| Guilty feelings (% endorsement) | 24.65% | 47.49% | 41.94% | 46.29% | 47.87% | 57.43% |

| | **Control group** | **Whole depression group** | **Cluster 1** | **Cluster 2** | **Cluster 3** | **Cluster 4** |
|---|---|---|---|---|---|---|
| Risk taking (% endorsement) | 25.83% | 28.60% | 28.63% | 30.81% | 26.64% | 30.07% |
| Depressed mood in last 2 weeks (median frequency) | Not at all | Not at all | Not at all | Not at all | Not at all | Several days |
| Unenthusiasm / disinterest in last 2 weeks (median frequency) | Not at all | Not at all | Not at all | Not at all | Not at all | Not at all |
| Tenseness / restlessness in last 2 weeks (median frequency) | Not at all | Not at all | Not at all | Not at all | Not at all | Not at all |
| Tiredness / lethargy in last 2 weeks (median frequency) | Not at all | Several days | Several days | Several days | Several days | Several days |

**Figures S3A-G. Visualizations of response distributions for nominal variables that significantly differed between depression clusters (data labels have been removed for values < 2%).** A: Milk type mainly used. B: Spread type mainly used. C: Bread type mainly eaten. D: Cereal type mainly eaten. E: Major dietary changes in the last 5 years. F: Alcohol consumption with meals. G: Smoking status.

**(A) Milk Type Mainly Used**

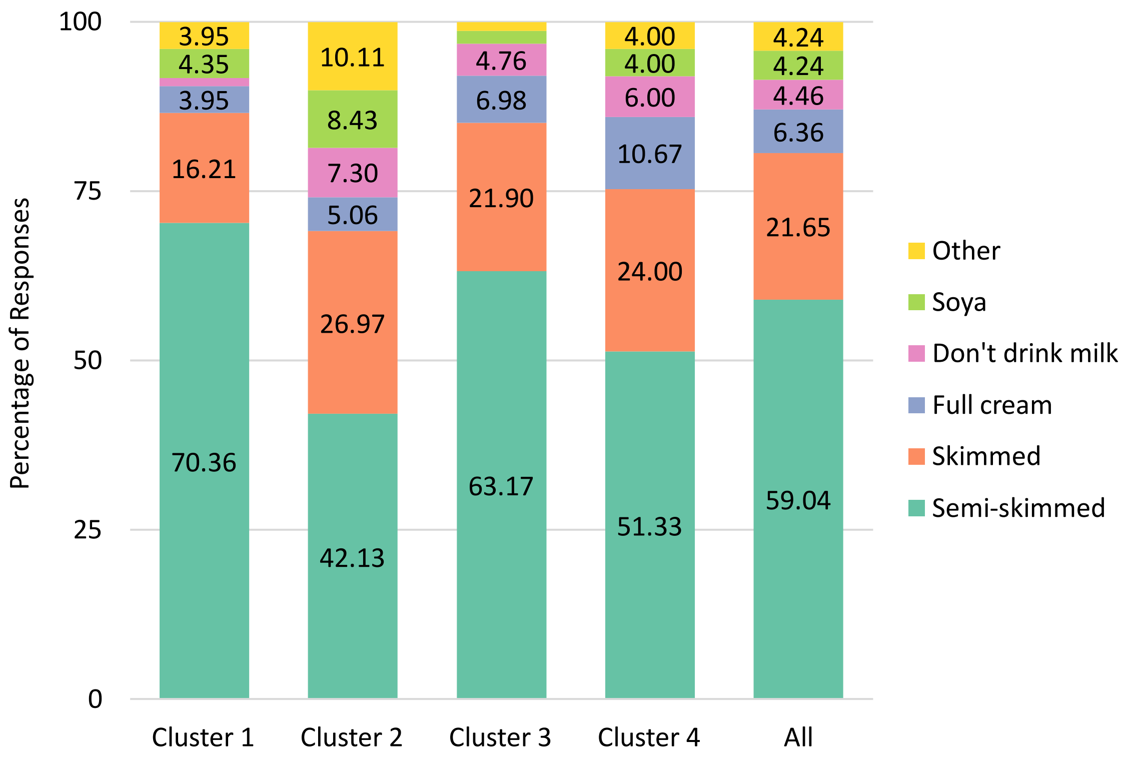

**(B) Spread Type Mainly Used (Butter or Butter Alternative)**

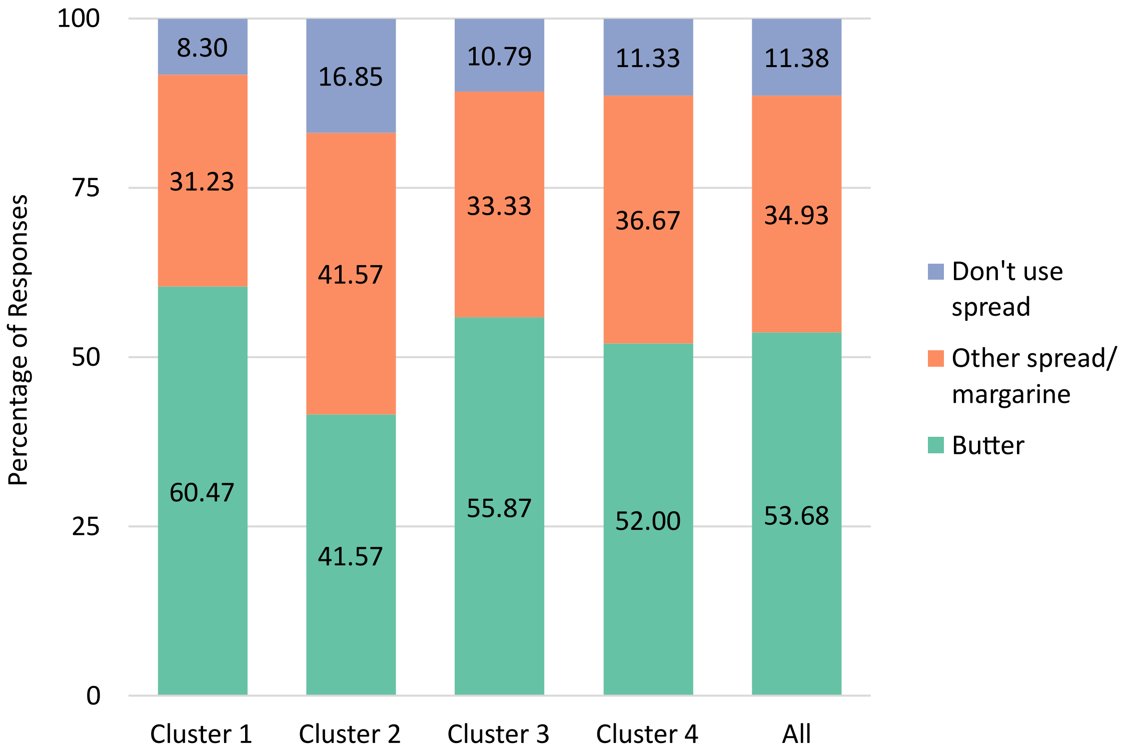

**(C) Bread Type Mainly Eaten**

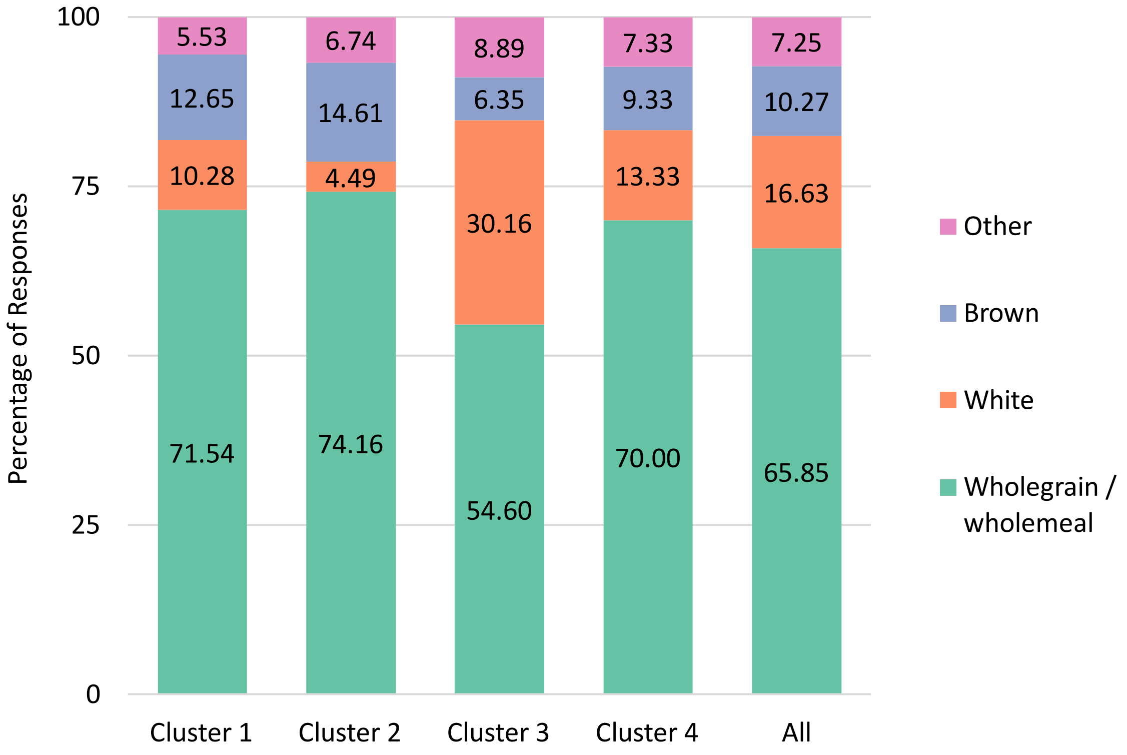

**(D) Cereal Type Mainly Eaten**

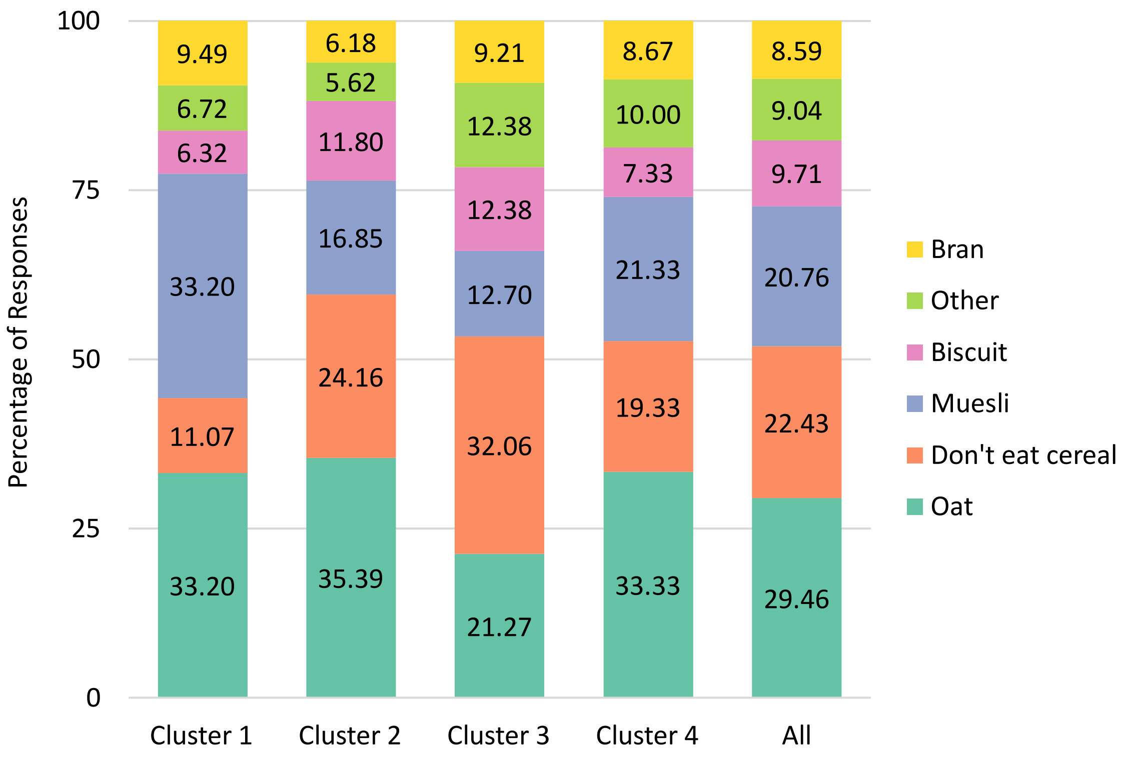

**(E) Major Dietary Changes in the Last 5 Years**

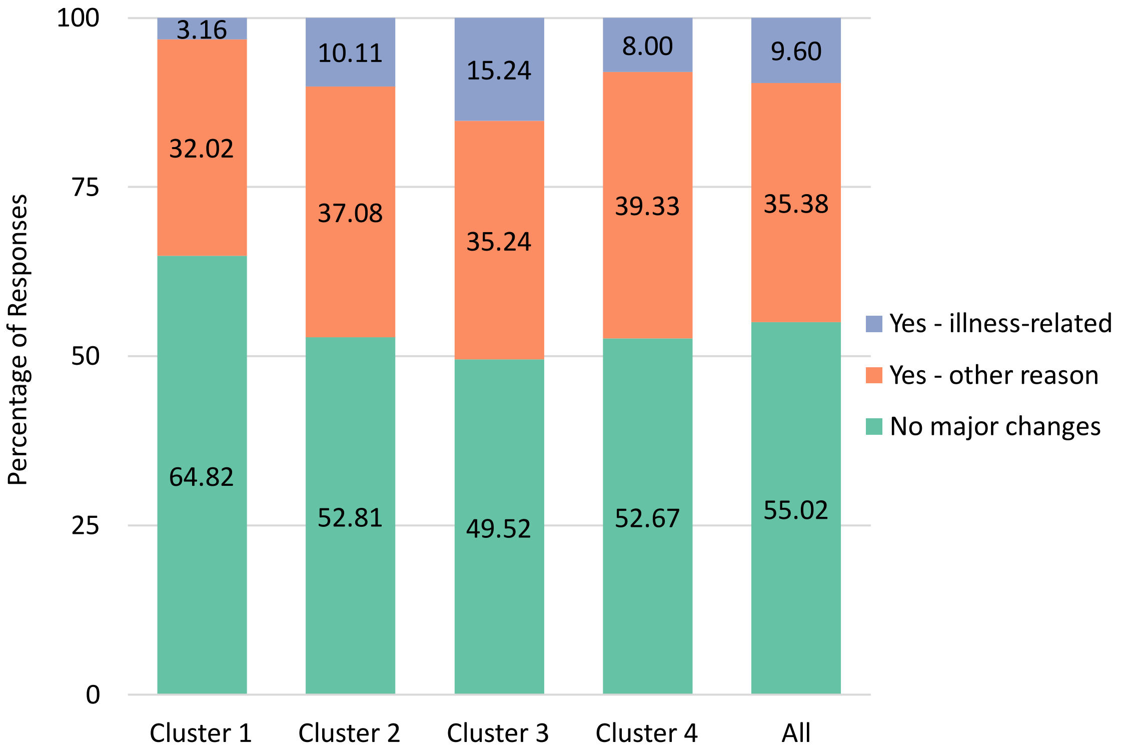

**(F) Alcohol Usually Taken With Meals**

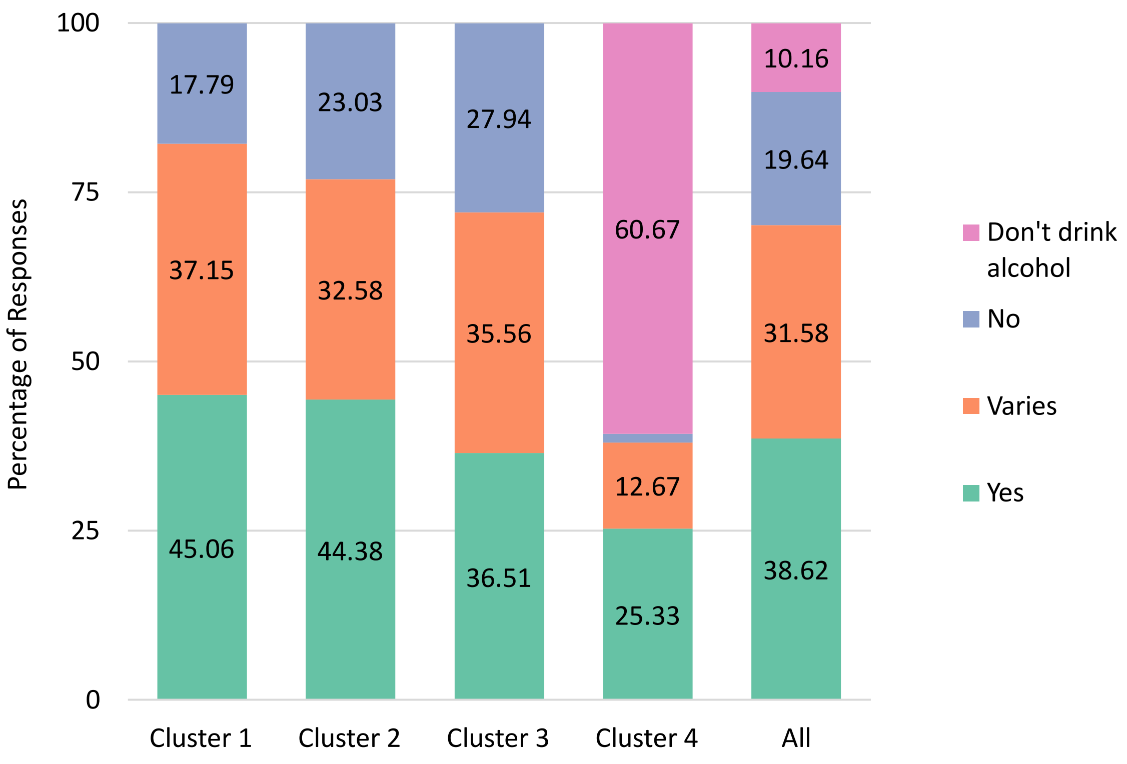

**(G) Smoking Status**

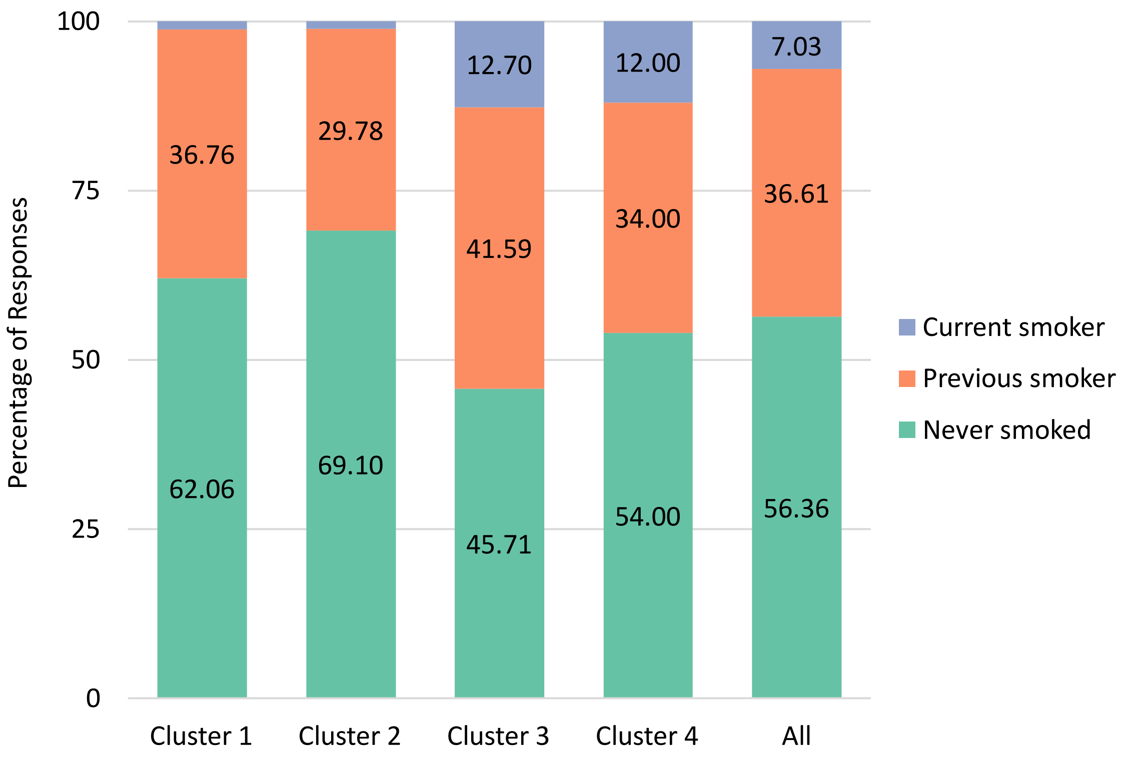

### 5. HYDRA Clustering Results Without Sex as a Covariate

Given the higher percentage of females in the depression group compared to the non-psychiatric reference group, our main analysis included sex (along with age) as a covariate in the HYDRA model. However, recognizing the potential clinical relevance of sex differences, we also conducted the following supplemental analysis without sex as a covariate. In this analysis, HYDRA identified four clusters within the depression group (ARI = 0.433; n = 265, 237, 163, and 144 for Clusters 1-4, respectively). Cluster differentiation was driven largely by the same features as in the main analysis, except for four variables that significantly differed between clusters here but not in the main analysis (non-oily fish intake, tea consumption, frequency of visits from friends/family, and heavy DIY activities) and one variable that significantly differed in the main analysis but not in this one (frequency of strenuous exercise). Additionally, the clusters differed in most of the same mood-related symptoms (except for guilt, which was not significant here) as well as in area deprivation and education. Notably, sex distribution varied across clusters, with Cluster 1 having the highest proportion of males (42%, compared to 34% in the depression group as a whole) along with the most severe psychopathology and unhealthy diet/lifestyle patterns.

**Table S5. Cluster differences in the HYDRA input features among adults with a history of depression (sex covariate removed from the HYDRA model).** Asterisks (*) indicate effects that remained significant after correcting for multiple comparisons ($P_{FDR} < .05$). Variable types and test statistics are indicated by superscript letters.

| | Cluster 1 | Cluster 2 | Cluster 3 | Cluster 4 | Statistic | df | P |
|---|---|---|---|---|---|---|---|
| HYDRA COVARIATES | | | | | | | |
| Age[a] (years) | 63.47 | 63.30 | 62.91 | 62.30 | 0.862 | 3, 892 | .460 |
| HYDRA INPUT FEATURES | | | | | | | |
| Diet | | | | | | | |
| Beef intake[c] | 41.02% | 39.16% | 18.18% | 32.72% | 29.048 | 3 | <.001* |
| Lamb/mutton intake[c] | 19.66% | 19.39% | 4.55% | 9.88% | 27.648 | 3 | <.001* |
| Pork intake[c] | 32.54% | 19.01% | 8.52% | 20.99% | 39.478 | 3 | <.001* |
| Poultry intake[c] | 0.00% | 0.00% | 0.00% | 0.00% | - | - | - |
| Processed meat intake[c] | 31.53% | 28.90% | 7.39% | 27.78% | 38.225 | 3 | <.001* |
| Non-oily fish intake[c] | 10.17% | 17.49% | 9.66% | 16.67% | 10.062 | 3 | .018* |
| Oily fish intake[c] | 15.25% | 20.91% | 17.05% | 25.31% | 7.910 | 3 | .048 |
| Cheese intake[c] | 13.22% | 11.79% | 25.00% | 22.22% | 19.359 | 3 | <.001* |
| Milk type mainly used[d] | Semi-skimmed 61.69% | Semi-skimmed 72.24% | Semi-skimmed 42.61% | Semi-skimmed 50.62% | 91.247 | 15 | <.001* |

| | Cluster 1 | Cluster 2 | Cluster 3 | Cluster 4 | Statistic | df | P |
|---|---|---|---|---|---|---|---|
| Spread type mainly used[d] | Butter 56.27% | Butter 61.22% | Butter or other/ margarine 84.09% (tied) | Butter 49.38% | 18.514 | 6 | .005* |
| Dried fruit intake[c] | 30.51% | 53.23% | 60.23% | 41.98% | 49.299 | 3 | <.001* |
| Fresh fruit intake[c] | 21.02% | 42.97% | 37.50% | 35.80% | 32.919 | 3 | <.001* |
| Cooked vegetable intake[c] | 18.64% | 23.95% | 34.09% | 24.69% | 14.320 | 3 | .003* |
| Salad / raw vegetable intake[c] | 23.05% | 47.53% | 37.50% | 23.46% | 46.486 | 3 | <.001* |
| Bread intake[c] | 53.90% | 46.77% | 43.75% | 45.68% | 5.854 | 3 | .119 |
| Bread type mainly eaten[d] | Wholegrain 54.92% | Wholegrain 69.20% | Wholegrain 74.43% | Wholegrain 70.99% | 99.354 | 9 | <.001* |
| Cereal intake[c] | 35.93% | 50.57% | 46.02% | 46.91% | 13.224 | 3 | .004* |
| Cereal type mainly eaten[d] | Don't eat cereal 31.19% | Muesli 31.94% | Oat 34.09% | Oat 34.57% | 70.487 | 15 | <.001* |
| Tea intake[c] | 41.69% | 51.71% | 39.20% | 50.00% | 9.937 | 3 | .019* |
| Water intake[c] | 34.58% | 40.68% | 62.50% | 36.42% | 39.234 | 3 | <.001* |
| Consume sugar[b] | 19.32% | 20.91% | 19.89% | 19.14% | 0.290 | 3 | .962 |
| Added salt intake[c] | 48.81% | 44.49% | 27.84% | 41.98% | 20.736 | 3 | <.001* |
| Major diet changes in past 5 years[d] | No 52.20% | No 60.08% | No 53.41% | No 53.70% | 31.136 | 6 | <.001* |
| Weekly variation in diet[c] | 9.49% | 9.13% | 7.95% | 12.96% | 2.693 | 3 | .441 |
| Nutritional supplements | | | | | | | |
| Fish oil[b] | 20.34% | 23.57% | 26.70% | 19.14% | 3.805 | 3 | .283 |
| Glucosamine[b] | 4.07% | 12.55% | 18.75% | 12.35% | 26.349 | 3 | <.001* |
| Vitamin D[b] | 15.93% | 20.53% | 18.18% | 19.75% | 2.195 | 3 | .533 |
| Multivitamins +/- minerals[b] | 24.41% | 14.83% | 32.39% | 27.16% | 19.966 | 3 | <.001* |
| Physical exercise | | | | | | | |
| Heavy DIY (last 4 weeks)[c] | 37.29% | 39.92% | 41.48% | 25.31% | 11.996 | 3 | .007* |
| Light DIY (last 4 weeks)[c] | 30.51% | 50.57% | 31.25% | 35.19% | 28.656 | 3 | <.001* |

| | Cluster 1 | Cluster 2 | Cluster 3 | Cluster 4 | Statistic | df | P |
|---|---|---|---|---|---|---|---|
| Strenuous sports (last 4 weeks)[c] | 5.08% | 10.27% | 6.25% | 6.79% | 5.994 | 3 | .112 |
| Walking for pleasure (last 4 weeks)[c] | 31.53% | 51.33% | 42.05% | 42.59% | 22.672 | 3 | <.001* |
| Other exercise (last 4 weeks)[c] | 19.32% | 47.53% | 56.25% | 49.38% | 83.843 | 3 | <.001* |
| Weekly sports club / gym participation[b] | 14.58% | 42.21% | 30.68% | 32.10% | 53.059 | 3 | <.001* |
| Alcohol use and smoking | | | | | | | |
| Alcohol intake frequency[c] | 43.05% | 51.71% | 38.64% | 23.46% | 33.945 | 3 | <.001* |
| Alcohol usually taken with meals[d] | Varies 37.97% | Yes 46.77% | Yes 44.32% | Don't drink 55.56% | 470.675 | 9 | <.001* |
| Smoking status[d] | Never 46.10% | Never 61.60% | Never 67.61% | Never 54.32% | 54.516 | 6 | <.001* |
| Social activities / relationships | | | | | | | |
| Pub or social club[b] | 24.07% | 31.56% | 22.73% | 10.49% | 24.819 | 3 | <.001* |
| Religious group[b] | 10.51% | 15.21% | 16.48% | 25.93% | 18.776 | 3 | <.001* |
| Other weekly group activity[b] | 26.44% | 43.73% | 56.82% | 27.16% | 55.088 | 3 | <.001* |
| No regular social activities[b] | 44.41% | 16.35% | 21.59% | 26.54% | 60.161 | 3 | <.001* |
| Frequency of friend/ family visits[c] | 43.39% | 54.37% | 40.34% | 42.59% | 11.256 | 3 | .010* |
| Frequency of being able to confide[c] | 35.93% | 51.33% | 44.32% | 41.36% | 13.754 | 3 | .003* |
| Often feels lonely[b] | 34.92% | 17.49% | 34.66% | 29.63% | 24.788 | 3 | <.001* |
| Outdoor exposure | | | | | | | |
| Time outdoors in the summer[c] | 39.32% | 47.15% | 57.39% | 41.36% | 15.975 | 3 | .001* |
| Time outdoors in the winter[c] | 35.25% | 48.67% | 57.95% | 41.98% | 25.292 | 3 | <.001* |
| Use of electronics / screentime | | | | | | | |
| Length of mobile use[c] | 0.00% | 0.00% | 0.00% | 0.00% | - | - | - |

| | Cluster 1 | Cluster 2 | Cluster 3 | Cluster 4 | Statistic | df | P |
|---|---|---|---|---|---|---|---|
| Weekly mobile use[c] | 39.66% | 36.50% | 47.16% | 36.42% | 5.944 | 3 | .114 |
| Plays computer games[b] | 25.76% | 20.15% | 17.61% | 22.22% | 4.947 | 3 | .176 |
| Time spent using a computer[c] | 47.46% | 31.94% | 39.20% | 40.12% | 14.008 | 3 | .003* |
| Time spent watching TV[c] | 43.05% | 34.98% | 30.11% | 33.95% | 9.276 | 3 | .026* |
| Sleep quality | | | | | | | |
| Sleep duration[a] (hours) | 7.15 | 7.29 | 7.24 | 7.38 | 1.368 | 3, 892 | .251 |
| Insomnia[c] | 43.73% | 32.70% | 36.36% | 43.21% | 8.885 | 3 | .031* |
| Physical health | | | | | | | |
| BMI[a] | 28.59 | 26.60 | 26.57 | 27.39 | 12.267 | 3, 892 | <.001* |
| Grip strength[a] | 28.14 | 27.47 | 25.93 | 25.47 | 3.545 | 3, 892 | .014* |
| VARIABLES NOT USED IN HYDRA | | | | | | | |
| Sex[b] (male) | 42.03% | 33.46% | 26.14% | 29.01% | 15.156 | 3 | .002* |
| G-brainAGE[a] | 0.30 | 0.17 | 0.52 | 0.34 | 0.198 | 3, 892 | .897 |
| Environment / socioeconomic status (SES) | | | | | | | |
| Townsend deprivation index[a] | -0.92 | -2.09 | -1.27 | -1.59 | 7.719 | 3, 891 | <.001* |
| Total household income[c] | 45.66% | 49.37% | 39.26% | 46.53% | 4.044 | 3 | .257 |
| Paid / self-employed[b] | 43.73% | 36.88% | 35.23% | 37.65% | 4.446 | 3 | .217 |
| Retired[b] | 54.24% | 58.56% | 56.82% | 51.23% | 2.501 | 3 | .475 |
| College or university degree[b] | 35.93% | 46.39% | 53.98% | 45.06% | 15.557 | 3 | .001* |
| Mental health | | | | | | | |
| Mood swings[b] | 67.59% | 67.44% | 69.14% | 72.50% | 1.448 | 3 | .694 |
| Miserableness[b] | 68.40% | 66.54% | 68.21% | 71.43% | 1.102 | 3 | .777 |
| Irritability[b] | 43.97% | 33.60% | 40.85% | 39.10% | 6.187 | 3 | .103 |
| Sensitivity / hurt feelings[b] | 68.42% | 63.42% | 70.11% | 67.31% | 2.517 | 3 | .472 |
| Fed-up feelings[b] | 62.12% | 48.08% | 58.38% | 57.23% | 11.538 | 3 | .009* |
| Nervous feelings[b] | 37.63% | 34.23% | 31.36% | 37.66% | 2.315 | 3 | .510 |

| | Cluster 1 | Cluster 2 | Cluster 3 | Cluster 4 | Statistic | df | P |
|---|---|---|---|---|---|---|---|
| Worrier / anxious feelings[b] | 70.69% | 70.27% | 61.85% | 70.89% | 4.961 | 3 | .175 |
| Tense / highly strung[b] | 26.64% | 25.98% | 33.73% | 31.85% | 4.316 | 3 | .229 |
| Worry too long after embarrassment[b] | 62.90% | 62.00% | 65.06% | 66.88% | 1.209 | 3 | .751 |
| Suffer from nerves[b] | 41.43% | 35.48% | 33.53% | 40.94% | 4.051 | 3 | .256 |
| Guilty feelings[b] | 47.02% | 43.02% | 47.09% | 55.90% | 6.668 | 3 | .083 |
| Risk taking[b] | 27.46% | 27.13% | 32.35% | 29.03% | 1.638 | 3 | .651 |
| Depressed mood in last 2 weeks[c] | 47.12% | 40.16% | 45.09% | 47.37% | 3.183 | 3 | .364 |
| Unenthusiasm / disinterest in last 2 weeks[c] | 46.64% | 31.01% | 37.57% | 39.74% | 14.065 | 3 | .003* |
| Tenseness / restlessness in last 2 weeks[c] | 45.67% | 44.09% | 43.79% | 40.99% | 0.924 | 3 | .820 |
| Tiredness / lethargy in last 2 weeks[c] | 29.97% | 15.69% | 20.00% | 24.68% | 16.769 | 3 | .001* |

[a]Continuous variable; values are means and *F* test statistic (one-way analysis of variance; ANOVA).

[b]Dichotomous variable; values are percentage who endorse the measure and $\chi^2$ test statistic (test for relationship between cluster and response).

[c]Ordinal variable; values are percentage above the pooled median and $\chi^2$ test statistic (test for relationship between cluster and percentage above the median).

[d]Nominal variable; values are categories with the highest percentage of endorsement and $\chi^2$ test statistic (test for relationship between cluster and response distributions).

$*P_{(PDR)} < .05$

**Figure S4. Visualization of HYDRA-based profiles of the clusters of individuals with a depression history (sex covariate removed from the HYDRA model).** Values are group summary measures (e.g., mean) that have been scaled for ease of comparison to range from 1 (cluster with the lowest value) to 3 (cluster with the highest value) for variables with significant group differences ($P_{FDR} < .05$). Variables comprising composite measures are: (1) Whole grains (intake of whole grain bread, muesli cereal, and oat cereal), (2) Fruits & vegetables (intake of fresh fruit, dry fruit, salad/raw vegetables, and cooked vegetables), (3) Soya/other milk/none (preference for soya milk, other milk, or no milk), (4) Supplements (glucosamine and multivitamins), (5) Red meat (intake of pork, beef, and lamb), (6) Full cream/semi-skim milk (preference for full cream or semi-skimmed milk), (7) Exercise (gym participation, heavy DIY, light DIY, walking for pleasure, and other exercise), (8) Physical health (low BMI and high grip strength), (9) Social support (low loneliness, high ability to confide in others, and frequent visits from family/friends), (10) Outdoor exposure (time spent outdoors in winter & in summer), and (11) Screentime (TV and computer use).

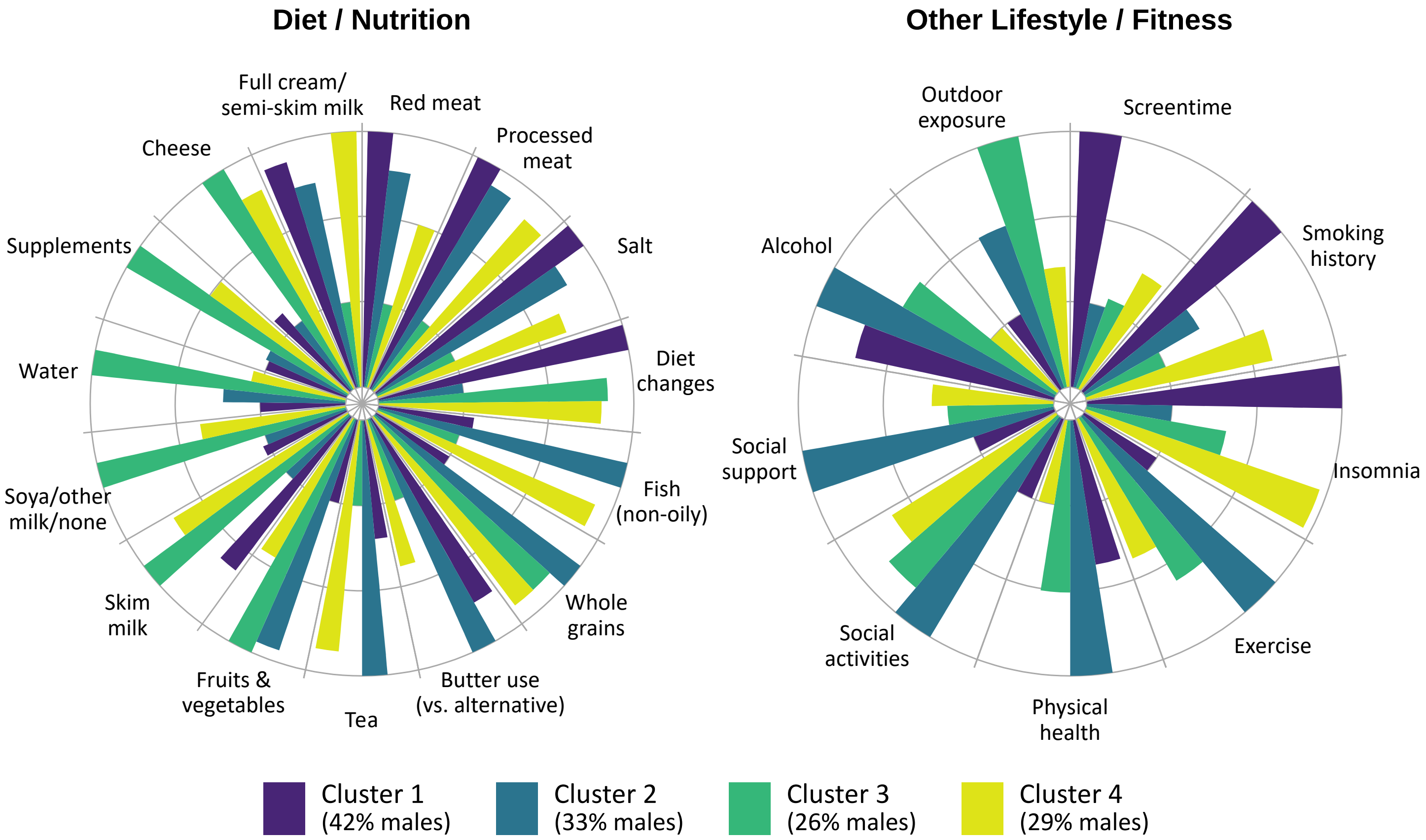

**Figure S5. Profiles distinguishing depression clusters with respect to socioeconomic indicators and psychopathology ratings (sex covariate removed from the HYDRA model).** Values are group summary measures (e.g., mean) that have been scaled for ease of comparison to range from 1 (cluster with the lowest value) to 3 (cluster with the highest value) for each variable with significant group differences ($P_{FDR} < .05$).

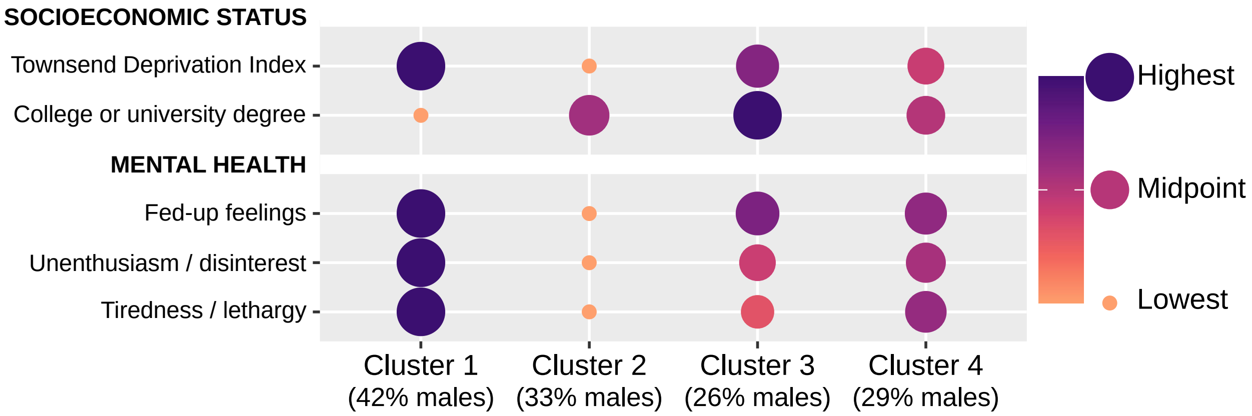

### 6. References

Varol, E., Sotiras, A., & Davatzikos, C. (2017). HYDRA: Revealing heterogeneity of imaging and genetic patterns through a multiple max-margin discriminative analysis framework. *NeuroImage*, *145*(Pt B), 346–364. https://doi.org/10.1016/J.NEUROIMAGE.2016.02.041
